# Air Pollution, Health, and Economics: Evaluating the Impact of WHO targets and Guideline Values on Mortality and Morbidity in Low- and Middle-Income Countries

**DOI:** 10.64898/2026.03.27.26349502

**Authors:** Annalan M D Navaratnam, Tom R P Bishop, Lambed Tatah, Harry Williams, Joseph V. Spadaro, Haneen Khreis

**Author notes:** Joint correspondence to: Dr Annalan M D Navaratnam, <u>01223 769200</u> Dr Haneen Khreis;, <u>01223 769200</u>.

## Abstract

**Background:** Ambient air pollution is a leading global health risk and disproportionately affects populations of Low- and Middle-Income Countries (LMICs). In 2021, WHO revised its Air Quality Guidelines (AQG), lowering recommended annual limits for Particulate Matter 2.5 (PM_2.5_) and Nitrogen Dioxide (NO_2_). We estimated the potential health and economic impacts of achieving WHO Interim Target 3 (IT3) and AQG concentrations across LMICs.

**Methods:** We conducted a health impact assessment across 136 LMICs to quantify one-year changes in all-cause and cause-specific mortality (chronic obstructive pulmonary disease [COPD], ischaemic heart disease [IHD], and stroke) and disease incidence (COPD, dementia, IHD, and stroke) under WHO IT3 and AQG counterfactual scenarios for PM_2.5_ and NO_2_. Concentration-response functions were applied at 1km x 1km resolution. Economic welfare impacts of mortality risk reductions were estimated using country-adjusted values of a statistical life (VSL, Int$ PPP-adjusted 2021). Direct medical and productivity-related costs associated with incident cases were estimated using a cost-of-illness (COI) framework. Uncertainty intervals (UI) reflect uncertainty in concentration-response functions.

**Results:** Attainment of WHO IT3 and AQG concentrations for PM_2.5_ was associated with an estimated 16.04% reduction (6.58million, UI: 6.10-7.07million) and 22.97% reduction (9.43million, UI: 8.75-10.11million) in annual deaths, respectively. Corresponding VSL-based estimates of deaths averted were Int$5.5 trillion (7.0% of aggregate LMIC GDP) and Int$8.4 trillion (10.6% of GDP), respectively. For NO_2_, IT3 and AQG scenarios were associated with estimated reductions of approximately 1.06% (approximately 435,000 deaths, UI: 388,000-483,000) and 2.79% (435,000 deaths; UI: 388,000-483,000), yielding gains of Int$0.6 trillion (0.7% of GDP) and Int$1.5 trillion (1.9% of GDP). Disease-specific mortality reductions were most prominent for IHD and stroke in Asia and Africa. Under the PM_2.5_ AQG scenario, an estimated 2.82million (1.67-2.97) COPD, 1.10million (0.83-1.37) dementia, 7.3million (6.41-8.19) IHD, and 2.3million (2.19-2.41) stroke cases could be delayed or averted in one year. Associated reductions in direct medical and productivity-related costs were greatest for IHD, COPD, and stroke. NO_2_-related morbidity reductions were smaller across all outcomes. All estimates represent one-year changes in risk relative to counterfactual exposure and may reflect delayed rather than permanently avoided events.

**Discussion:** Achieving both WHO IT3 and AQG values in LMICs could yield substantial reductions in premature mortality and disease incidence, particularly for cardiovascular and respiratory conditions, alongside large, monetised welfare gains from reduced mortality risk. These findings underscore the considerable societal value of air quality improvements and support accelerated action toward meeting WHO guideline levels in regions bearing the highest pollution burden.

## Introduction

Ambient (i.e., outdoor) air pollution is the largest environmental threat to human health worldwide, adversely impacting workforce productivity and overall economic activity.^1,2^ Due to growing evidence of its potency, the World Health Organisation lowered the annual average air quality guideline values for several pollutants, including particulate matter 2.5 (PM_2.5_) from 10 to 5 μg/m^3^ and nitrogen dioxide (NO_2_) from 40 to 10 μg/m^3.3^ An estimated 0.18% of the global land area and 0.001% of the global population had annual exposure below the updated PM_2.5_ annual mean concentration threshold in 2019.^4^ The greatest exposure is seen in low- and middle-income countries (LMIC), where air quality has generally deteriorated in line with large-scale urbanisation, economic development, and increasing motorisation that has mostly relied on burning of fossil fuels.^5^

Most of the mortality burden attributable to outdoor air pollution is related to cardiometabolic conditions, particularly ischaemic heart disease (IHD) and stroke.^6^ IHD, stroke, and chronic obstructive pulmonary disease (COPD) are ranked as the top leading causes of deaths globally for the past two decades (excluding COVID19), with the highest burden in LMIC.^7^ These have also been the leading three causes of deaths and DALYs attributable to ambient PM_2.5_.^8^ In LMICs it is likely that access to healthcare is limited, and therefore higher rates of morbidity and mortality are resulting in significant economic and health costs.^9^ IHD and stroke mortality in low-income countries has been reported to be more than twice and three times that of mortality in high-income countries (HICs), respectively.^10^ With a rapidly aging population globally, IHD and stroke are likely to increase in incidence incurring larger economic and human costs.^11^ In addition to these diseases, dementia (i.e., Alzheimer’s Dementia and other dementias) is projected to rapidly increase in prevalence from 51.2 million in 2021 to 152.8 million by 2050, with the largest increases in LMIC.^12^ As well as having a significant impact to society, the affected individual and their caregivers, dementia has a significant economic burden with estimates as high as $2.8 trillion (US$ 2020 constant) in 2019 and projected to be $16.9 trillion (95% Confidence Interval (CI) $11.3 – 27.3 trillion) by 2050 globally.^13^

Whilst the Lancet Commission on Dementia Prevention, Intervention and Care 2024 recognised air pollution as a modifiable risk factor, air pollution is not recognised as a risk factor for cardiovascular disease in WHO’s Non-Communicable Disease action plan, despite the plan outlining the need for prevention and treatment options to reduce premature mortality from NCDs.^14,15^ The majority of studies on health and economic impacts of ambient air pollution have been conducted in high-income countries in Europe and North America, many of which have the lowest national exposure levels.^3^ By 2050, 90% of urban population growth, i.e., movement from rural to urban settings in addition to actual growth, is predicted to occur in cities of LMIC in Asia and Africa.^16^ Therefore substantive evidence on human and economic cost is needed to guide policy decisions in LMIC settings to reduce air pollution, and subsequent disease outcomes. In the context of competing public health challenges, evidence may highlight how pollution reduction policies can have significant health and economic benefits. This will enable policy makers to make data-driven decisions that aim to improve the population health and reduce economic burden.

## Methods

### Overview

This study conducted a quantitative health impact assessment (HIA) to estimate the potential health and economic benefits in LMICs by reducing concentrations of two air pollutants, PM_2.5_ and NO_2_. LMIC was defined by the Organisation for Economic Co-operation and Development’s (OECD’s) list of 141 Official Development Assistance Country recipients (2024-2025 flow; Supplementary Table 1).^17^

Two counterfactual scenarios were modelled to estimate potential reductions from baseline in mortality and disease burden using WHO recommended annual mean pollutant concentrations from the 2021 WHO Global Air Quality Guidelines:

1. WHO Interim Target 3 (IT3): 15µg/m^3^ for PM_2.5_ and 20µg/m^3^ for NO_2_.
2. WHO Air Quality Guideline (AQG): 5µg/m^3^ for PM_2.5_ and 10µg/m^3^ for NO_2_.

The WHO AQG counterfactual scenario has been assumed as the theoretical minimum risk exposure level (TMREL), which is defined as the lowest point of the risk function and it represents the level of risk exposure that minimises disease risk at the population level.^18^ Therefore it can be assumed that the number of annual deaths or cases prevented by meeting the AQG concentrations also reflect mortality or disease incidence attributable to the pollutant.

### Population, Pollutant and Outcome Data

For this analysis, modelled population data were sourced from WorldPop’s constrained (UN adjusted) datasets, which provides high-resolution estimates of population distribution in 2019 at the 100m x 100m grid level within areas mapped as containing built settlements.^19^ Global data on PM_2.5_ (2022, approx. 1km x 1km resolution) and NO_2_ (2020, 50m x 50m resolution) annual average concentrations were sourced from Shen et al and Larkin et al, respectively.^20,21^ Although the WorldPop resolution is provided at 100m x 100m resolution, it was aggregated to approximately 1km x 1km to match the pollutant maps resolution, which was standardised to approximately 1km x 1km due to limitations of computational power.^22^ Baseline incidence and mortality data by age (i.e., five-year age bands), sex, and country, for COPD, IHD, and stroke were sourced from the Institute for Health Metrics and Evaluation (IHME) Global Burden of Disease (GBD) data for 2021.^1^ Baseline incidence for dementia was also sourced from IHME by age, sex, and country.^1^ Health outcomes were selected based on globally ranked causes of mortality as well as predicted increasing prevalence, which also aligned with causality determinations (Supplementary ‘Methods 1. Population, pollutant and outcome selection and justification’ and Supplementary Table 2 for data sources, resolution and year).

### Health Impact Assessment

We applied concentration-response functions (CRFs) derived from the epidemiological literature to estimate potential one-year changes in mortality and disease incidence attributable to hypothetical reductions in ambient air pollutant concentrations under WHO counterfactual scenarios (Supplementary Table 3).

Changes in mortality or disease incidence were estimated at a 1km x 1km grid-cell level and subsequently aggregated to the country level. For each pollutant, disease outcome, age group, and country, the change in annual cases or deaths were calculated as:

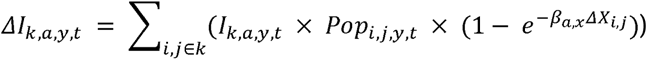

Where:

- *ΔI_k,a,y,t_* is the estimated change in annual incidence or mortality for country *k*, outcome *a*, age category *y* and sex *t*;
- *I_k,a,y,t_* is the baseline country-level incidence or mortality country *k,* outcome *a*, age category *y* and sex *t*;
- *Pop_i,j,y,t_* is the population size for age category *y* and sex *t* in grid cell *i,j*;
- *β_a,x_* is the outcome □ and pollutant *x* specific log-linear risk coefficient derived from the CRF;
- *Δx_i,j_*is the change in annual mean pollutant concentration in grid cell *i,j* between the baseline and counterfactual scenario.

Baseline incidence and mortality rates were obtained at the country level and applied uniformly across grid cells within each country, which population counts and pollution concentrations were spatially resolved at 1km x 1km resolution.

#### Application constraints and exposure limits

CRFs were applied only within the age range for which they were derived, excluding population groups under 60 years for dementia and under 30 for all other outcomes, by restricting the baseline incidence or mortality rates to the relevant age ranges. For grid cells in which baseline pollution concentration were already below the relevant counterfactual level, the relative risk was assumed to be 1, implying no change in risk under the scenario. Concentrations were truncated at the upper bound of the CRF exposure range and capped concentration difference was used in the impact calculations, to reduce the risk of overestimating health impacts in regions with relatively high pollutant concentrations and limited epidemiological evidence (Supplementary Table 3). ^23^

#### Uncertainty Characterisation

Uncertainty intervals (UI) were derived exclusively from the reported 95% confidence intervals (CI) of the CRFs by applying the lower and upper bounds of the risk coefficient (*β_a_*,) in the impact calculations. Uncertainty associated with pollutant concentration estimates, population data, or baseline incidence and mortality were not included. Consequently, the reported uncertainty intervals reflect epidemiological uncertainty only and should be interpreted as indicative ranges rather than probabilistic confidence intervals.

#### Economic Assessment

Valuation of mortality risk used the Value of Statistical Live (VSL) approach, adjusting the OECD 2005 valuation of $3 million (Purchasing Power Parity USD) by time (i.e., 2021 using consumer price inflation and Gross Domestic Income (GDP) per capita OECD countries) and geography (i.e., GDP per capita). The economic costs associated with disease-specific morbidity were estimated using cost-of-illness (COI) approach, for direct medical costs and indirect productivity losses associated with each case averted (Full methods in Supplementary Methods 2. Economic Assessment).

## Results

### Data

From the OECD Development Assistance Committee (DAC) list (141), five countries (Kosovo, Montserrat, Saint Helena, Wallis and Futuna, and (the State of) Palestine were excluded because health data was not available in the IHME database (see Supplementary Figure 1). The remaining 136 countries covered the continents of Africa (53), Asia (35), Americas (26), Oceania (14), and Europe (8), giving a total population of 2.97 billion (30 years and older; 50.2% female) and 0.67 billion (65 years and older; 52.4% female) people. The annual spatial mean concentration of PM_2.5_ and NO_2_ was 30.44 µg/m³ (Standard Deviation [SD] 18.06; range 0.6-266.26 µg/m³) and 6.39 µg/m³ (SD 5.98; range 0-66.45 µg/m³), respectively. The countries with the highest mean PM_2.5_ concentration, in descending order, were Iraq (68.91µg/m³), Bangladesh (65.66 µg/m³), Nigeria (62.32 µg/m³), Niger (61.55 µg/m³), Pakistan (58.30 µg/m³). For NO_2_ it was Lebanon (15.15 µg/m³), Jordan (13.77 µg/m³), China (13.33 µg/m³), Egypt (12.32 µg/m³), Armenia (11.60 µg/m³). At continent-level, Asia (29.69 µg/m³) and Africa (27.42 µg/m³) had the highest mean PM_2.5_ concentration and for NO_2_ it was Europe (8.66 µg/m³) and Asia (8.16 µg/m³). Oceania had the lowest mean concentration for PM_2.5_ (4.03 µg/m³) and NO_2_ (0.44 µg/m³). It was not possible to calculate health or economic impacts from NO_2_ reductions in nine countries, mostly in Oceania, because the concentration data were not available in these areas (Kiribati, Maldives, Marshall Islands, Mauritius, Nauru, Samoa, Tokelau, Tonga, and Tuvalu).

### Changes in Mortality and associated Monetary Value

#### Averted Premature Mortality

Across all LMIC, achieving the WHO IT3 and AQG guideline concentrations was associated with substantial estimated reductions in all-cause and disease-specific mortality. For PM_2.5_, attainment of WHO IT3 concentrations was associated with an estimated 16.04% reduction in annual deaths (6.58 million, UI: 6.10-7.07 million), while attainment of WHO AQG levels was associated with a 22.97% reduction (9.43 million, UI: 8.75-10.11 million) across 136 countries (Figure 1 and Table 1).

**Figure 1:**
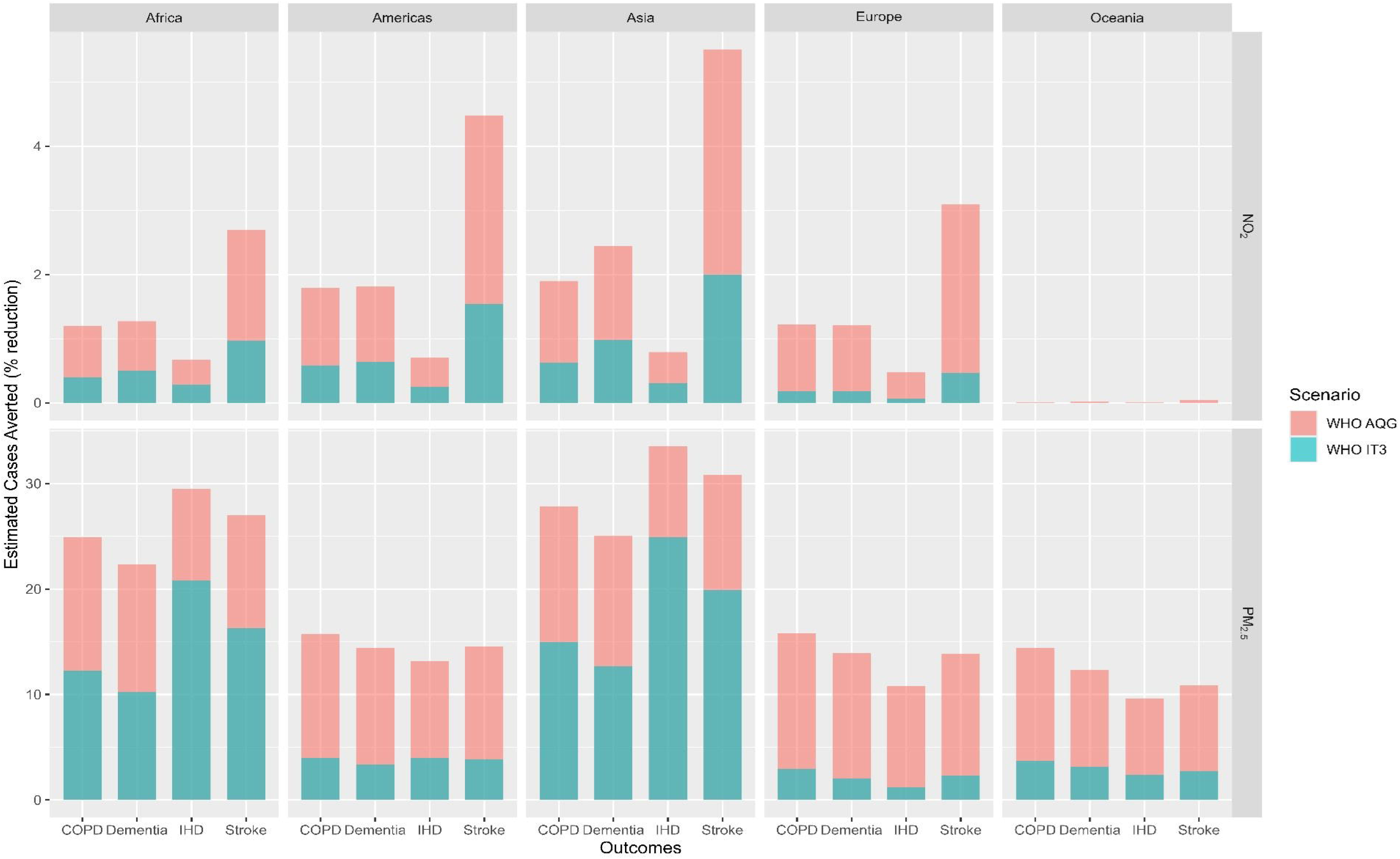
Estimated deaths averted in one-year from counterfactual scenarios (WHO IT3, and WHO AQG) as percentage reduction from mortality under ‘Current’ scenario (2021 IHME mortality estimates).

**Table 1:**
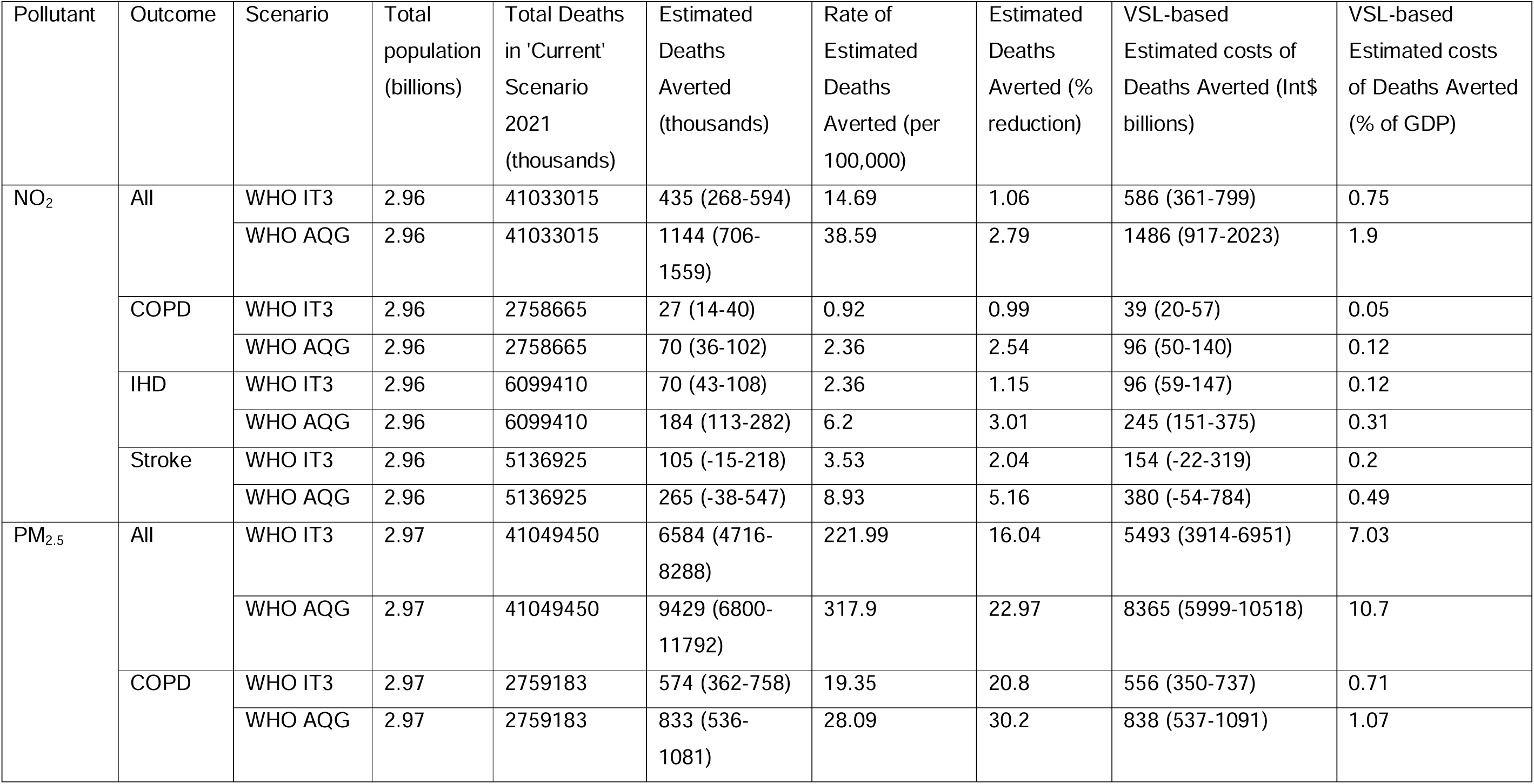
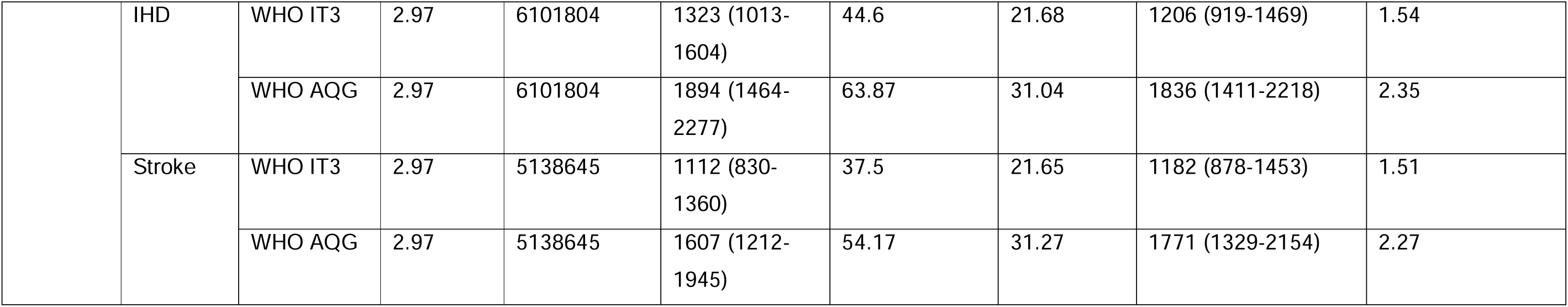
One-year change in all-cause and cause-specific mortality under hypothetical air pollution reduction scenarios and associated VSL-based economic welfare estimates of deaths averted, expressed relative to aggregate LMIC GDP. Total GDP for NO_2_ is Int$ 78,100.84 and PM_2.5_ Int$78,142.4 bill.

The largest disease-specific mortality reductions attributable to PM_2.5_ were observed for IHD and stroke. Under the IT3 scenario, IHD- and stroke-related mortality declined by 21.68% (1.32 million, UI: 1.23-1.41 million) and 21.65% (1.11 million, UI: 1.03-1.20 million), respectively. Under the AQG scenario, these reductions increased to 31.04% (1.89 million, UI: 1.77-2.02 million) for IHD and 31.27% (1.61 million, UI: 1.49-1.72 million) for stroke.

Reductions in COPD mortality were of similar relative magnitude (21.68% under IT3 and 31.04% under AQG), although absolute reductions were smaller, reflecting lower baseline mortality (574,000 [UI: 503,000-645,000] and 833,000 [UI: 503,000-645,000], respectively).

In contrast, achieving WHO IT3 and AQG levels for NO_2_ were associated with more modest reductions in annual mortality, corresponding to 1.06% (approximately 435,000 deaths, UI: 388,000-483,000) and 2.79% (1.14 million deaths; UI: 706,000-1.56 million), respectively (Figure 2 and Table 1). Although reductions in IHD and stroke mortality were also observed under NO_2_ scenarios, their magnitude was substantially smaller than those associated with PM_2.5_ (Figure 1 and Table 1). All mortality estimates reflect one-year change in mortality risk relative to the counterfactual and therefore represent potential delays in premature deaths rather than permanent avoidance over the life course.

**Figure 2:**
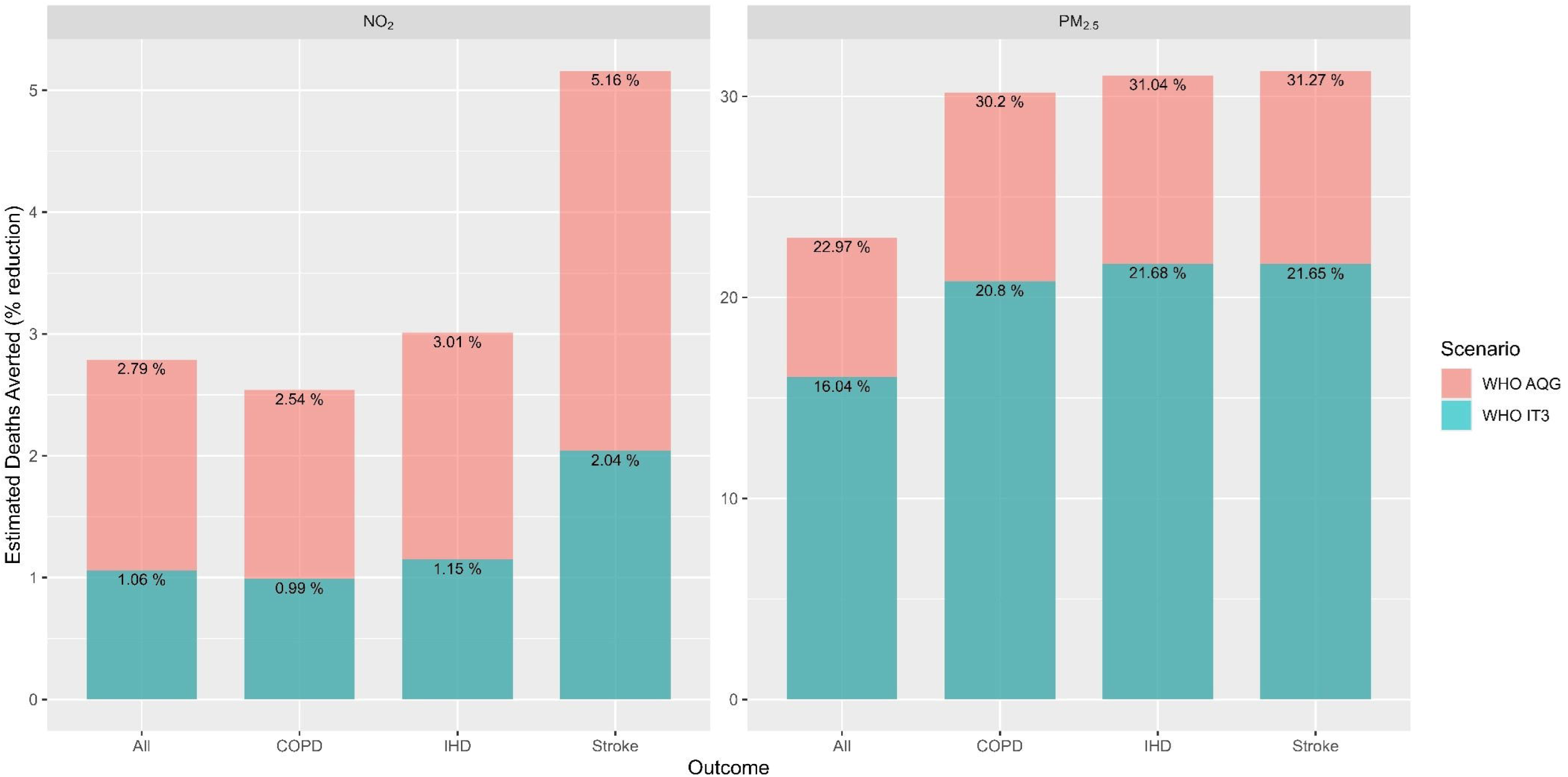
Estimated deaths averted in one-year from counterfactual scenarios (WHO IT3, and WHO AQG) by continent as percentage reduction from mortality under ‘Current’ scenario (2021 IHME mortality estimates).

##### Differences by geography and demography

Reductions in annual mortality under WHO IT3 and AQG scenarios were consistently larger for PM_2.5_ than for NO_2_ across all geographical regions (Figure 2). The largest absolute and relative reductions attributable to PM_2.5_ were observed in Asia (e.g., 19.2% under IT3 and 26.1% under AQG), reflecting high baseline exposures and population size. Relative reductions in Africa were also pronounced (Figure 2). China and India accounted for the largest absolute reductions in all-cause and disease-specific mortality across all pollutants and scenarios. However, the largest relative reductions for PM_2.5_ was highest in Bangladesh, Iraq, and Egypt for most outcomes, while COPD reductions were greatest in Iraq, Niger, and Bangladesh (Supplementary Figure 2). For NO_2_, Lebanon, Egypt, and Jordan consistently exhibited the largest relative mortality reductions across outcomes and scenarios (Supplementary Figure 2).

When disaggregated by sex, absolute and relative mortality reductions were broadly similar (Figure 3 and Supplementary Table 4). However, when stratified by both age and sex, larger absolute reductions were observed among males in younger age groups (40-59 years), regardless of pollutant, outcome, scenario (Supplementary Figures 4 and 5). This pattern likely reflects higher baseline mortality rates among younger males rather than differential exposure-response relationships.^24,25^

**Figure 3:**
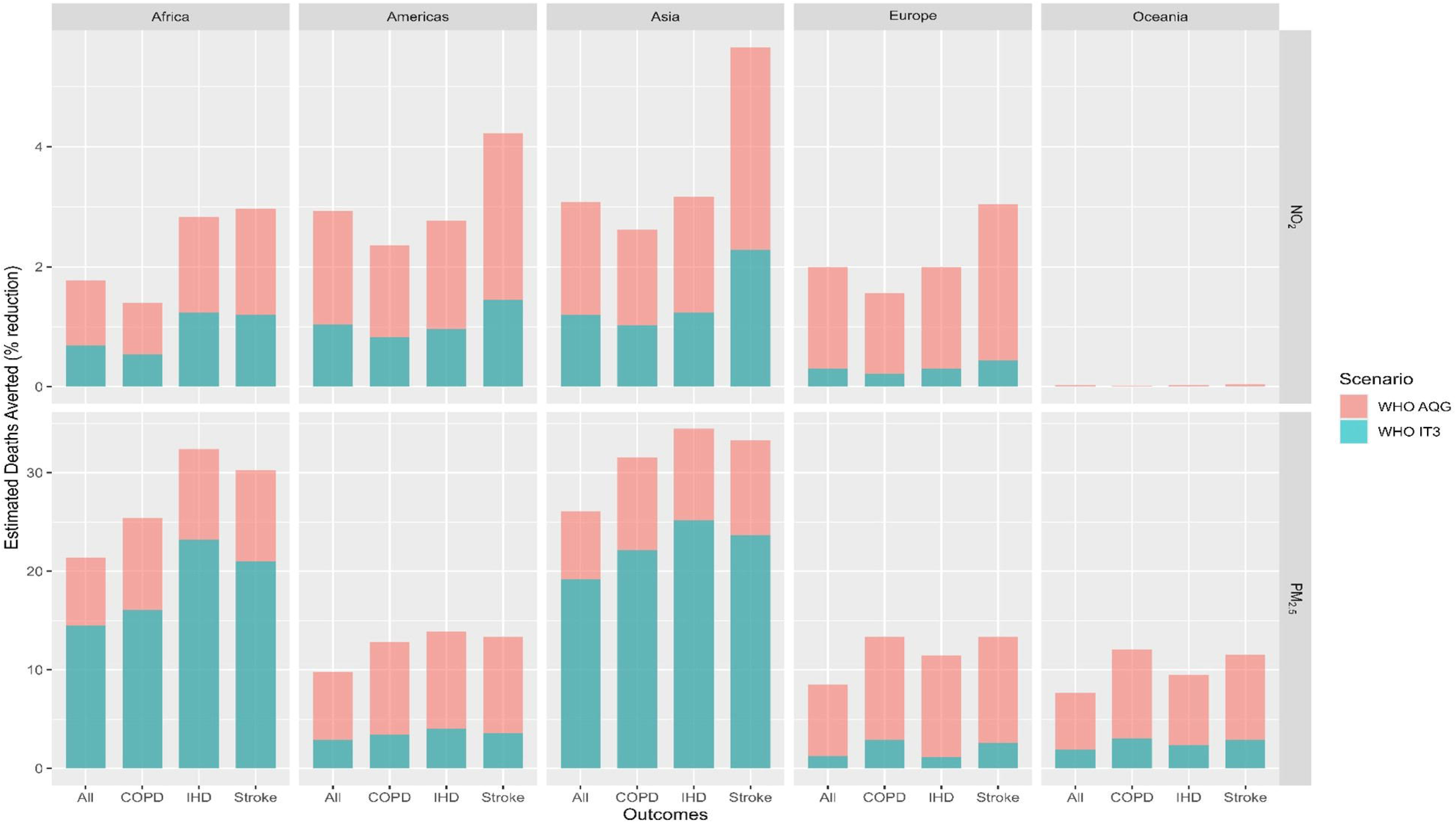
Estimated deaths averted in one-year from counterfactual scenarios (WHO IT3, and WHO AQG), stratified by sex, by continent as percentage reduction from mortality under ‘Current’ scenario (2021 IHME mortality estimates).

#### Economic Welfare Benefits of Averted Premature Mortality

When expressed in monetary terms, the monetised economic welfare benefits associated with one-year reductions in premature mortality risk from attaining WHO IT3 levels was an estimated Int$ 5.5 trillion (UI: 5.1-5.9 trillion) for PM_2.5_ and Int$ 0.6 trillion (0.5-0.7 trillion) for NO_2_, corresponding to 7.03% and 0.7% equivalent of the aggregate LMIC GDP (i.e., a market indicator), respectively (Table 1). Under the more stringent WHO AQG scenario, VSL-based estimates of deaths averted increased to Int$ 8.4 trillion (UI: 7.7-9.0 trillion) for PM_2.5_ and Int$ 1.5 trillion (UI: 1.3-1.7 trillion) for NO_2_, equivalent to 10.70% and 1.90% of aggregate LMIC GDP, respectively. These values represent monetised changes in population-level mortality risk rather than realised economic output.

Across causes of death, IHD and stroke accounted for the largest shares of VSL-based estimates associated with reduced mortality risk, with values of Int$ 1.84 trillion (UI: 1.70-1.97 trillion; equivalent to 2.35% of aggregate GDP) and Int$ 1.77 trillion (UI: 1.63-1.91 trillion; 2.27% of aggregate GDP), respectively. China exhibited the largest absolute welfare benefits from reduced mortality risk across all outcomes, pollutants, and scenarios, followed by India (except for NO_2_-based scenarios). When expressed relative to national GDP, China and India ranked among the top two countries only for COPD outcomes under PM_2.5_ reductions, while China ranked highest for COPD and stroke from NO_2_ reductions (Supplementary Figure 6 and 7 for PM_2.5_ and NO_2_, respectively).

### Averted Incidence, Years of Live with Disability and associated costs

#### Health Impact

Reductions in PM_2.5_ concentrations were associated with the largest estimated reduction in annual disease incidence across all outcomes and scenarios (Table 2). Under the WHO IT3 scenario, PM_2.5_ reductions were associated with a 13.51% reduction in COPD cases (1.45 million, UI: 1.37-1.53 million), 11.04% in dementia (524,000, UI: 389,000-659,000), 22.21% in IHD (5.25 million, UI:4.59-5.91 million), and 17.91% in stroke (1.43 million, UI: 1.36-1.5 million). Under the WHO AQG scenario, larger reductions were observed; 26.27% for COPD (2.82 million cases, UI: 1.67-2.97 million), 23.21% for dementia (1.10 million, UI: 0.83-1.37 million), 30.88% for IHD (7.3 million, UI: 6.41-8.19 million), and 28.82% for stroke (2.3 million, UI: 2.19-2.41 million).

**Table 2:** One-year change in incidence and years of live with disability (YLD) under hypothetical air pollution reduction scenarios and associated direct (i.e., medical) and indirect (i.e., labour productivity) costs of cases averted, expressed relative to aggregate LMIC GDP.

| Pollutant | Outcome | Scenario | Current incidence (thousands) | Current YLD (millions) | Cases averted | Cases averted (% reduction) | YLD averted (millions) | YLD averted (% reduction) | Direct Medical costs of cases averted (Int\$ mill) | Indirect costs to labour by YLD averted (Int\$ mill) | Total cost of cases averted (% of GDP) |
| --- | --- | --- | --- | --- | --- | --- | --- | --- | --- | --- | --- |
| NO<sub>2</sub> | COPD | WHO IT3 | 10732.31 | 3694.11 | 64092 (-218740-302300) | 0.6 | 20.60 (-70.33-97.15) | 0.56 | 76 (-260-360) | 457 (-1562-2156) | 0.001 |
|  |  | WHO AQG | 10732.31 | 3694.11 | 195037 (-687375-894888) | 1.82 | 63.12 (-22.23-28.97) | 1.71 | 227 (-800-1038) | 1349 (-4765-6181) | 0.002 |
|  | Dementia | WHO IT3 | 4743.527 | 2011.81 | 41453 (14200-67273) | 0.87 | 17.78 (6.09-28.85) | 0.88 | 394 (135-639) | 0 (0-0) | 0 |
|  |  | WHO AQG | 4743.527 | 2011.81 | 105701 (36282-171217) | 2.23 | 45.28 (15.54-73.34) | 2.25 | 995 (342-1612) | 0 (0-0) | 0.001 |
|  | IHD | WHO IT3 | 23633.25 | 956.84 | 69063 (50520-90428) | 0.29 | 2.90 (2.12-3.80) | 0.3 | 170 (124-223) | 67 (49-88) | 0 |
|  |  | WHO AQG | 23633.25 | 956.84 | 179111 (131054-234452) | 0.76 | 7.53 (5.51-9.86) | 0.79 | 432 (316-565) | 170 (125-223) | 0.001 |
|  | Stroke | WHO IT3 | 7971.854 | 3455.64 | 143470 (206596-74854) | 1.8 | 63.75 (91.79-33.26) | 1.84 | 475 (684-248) | 1286 (1852-671) | 0.002 |
|  |  | WHO AQG | 7971.854 | 3455.64 | 400782 (572944-210752) | 5.03 | 177.10 (253.15-93.14) | 5.12 | 1309 (1871-689) | 3481 (4973-1832) | 0.006 |
| PM<sub>2.5</sub> | COPD | WHO IT3 | 10735.39 | 3695.19 | 1450080 (1094583-1776111) | 13.51 | 522.09 (394.16-639.36) | 14.13 | 1193 (900-1461) | 9411 (7105-11527) | 0.013 |
|  |  | WHO AQG | 10735.39 | 3695.19 | 2820301 (2168497-3395393) | 26.27 | 994.78 (765.28-1197.01) | 26.92 | 2454 (1884-2958) | 18118 (13935-21808) | 0.026 |
|  | Dementia | WHO IT3 | 4745.651 | 2012.72 | 524009 (142181-849287) | 11.04 | 221.62 (60.13-359.20) | 11.01 | 3915 (1061-6349) | 0 (0-0) | 0.005 |
|  |  | WHO AQG | 4745.651 | 2012.72 | 1101618 (314033-1708772) | 23.21 | 466.56 (132.99-723.77) | 23.18 | 8615 (2449-13397) | 0 (0-0) | 0.011 |
|  | IHD | WHO IT3 | 23638.9 | 957.24 | 5250979 (2350066-7645340) | 22.21 | 190.93 (85.03-279.32) | 19.95 | 7738 (3406-11444) | 3665 (1631-5364) | 0.014 |
|  |  | WHO AQG | 23638.9 | 957.24 | 7300729 (3336110-10428542) | 30.88 | 275.07 (124.87-395.38) | 28.74 | 11804 (5294-17156) | 5333 (2418-7675) | 0.022 |
|  | Stroke | WHO IT3 | 7974.809 | 3457.37 | 1428199<br>(1123882-1704759) | 17.91 | 620.37 (488.12-740.60) | 17.94 | 3494 (2747-4175) | 11006<br>(8660-13138) | 0.019 |
|  |  | WHO AQG | 7974.809 | 3457.37 | 2298462<br>(1832270-2710571) | 28.82 | 998.94 (796.26-1178.13) | 28.89 | 5859 (4663-6920) | 17852<br>(14228-21058) | 0.03 |

Reductions in Years of Life with Disability followed a similar but not identical pattern. Under the PM_2.5_ AQG scenario, the largest relative YLD reductions were for stroke (28.89%), IHD (28.74%), COPD (26.92%) and dementia (23.18%). In contrast, NO_2_ reductions yielded smaller relative declines in disease incidence and YLD, with the largest relative reduction observed for stroke (5.03% and 5.12%, respectively) under the AQG scenario. All morbidity estimates reflect one-year changes in disease incidence and isolated to one disease and therefore represent delayed onset rather than permanent prevention nor prevention of any disease.

##### Differences by geography and demography

The spatial distribution of cases averted reflects regional differences in baseline exposure and population size, with the largest relative reductions observed in Asia and Africa for PM_2.5_. For NO_2_, the largest relative reductions were observed in Asia and the Americas, while impacts in Africa and Europe were more modest and negligible in Oceania (Figure 4). As observed for mortality, China and India accounted for the largest absolute reductions in disease incidence across pollutants and scenarios. However, the largest relative reductions for PM_2.5_ were COPD and stroke incidence in The Gambia, Benin, and Burundi and for IHD incidence were greatest in Iraq, Bangladesh, and Egypt (Supplementary Figure 8). For NO_2_, Lebanon, Egypt, and Jordan consistently exhibited the largest relative reductions across outcomes and scenarios (Supplementary Figure 9).

**Figure 4:**
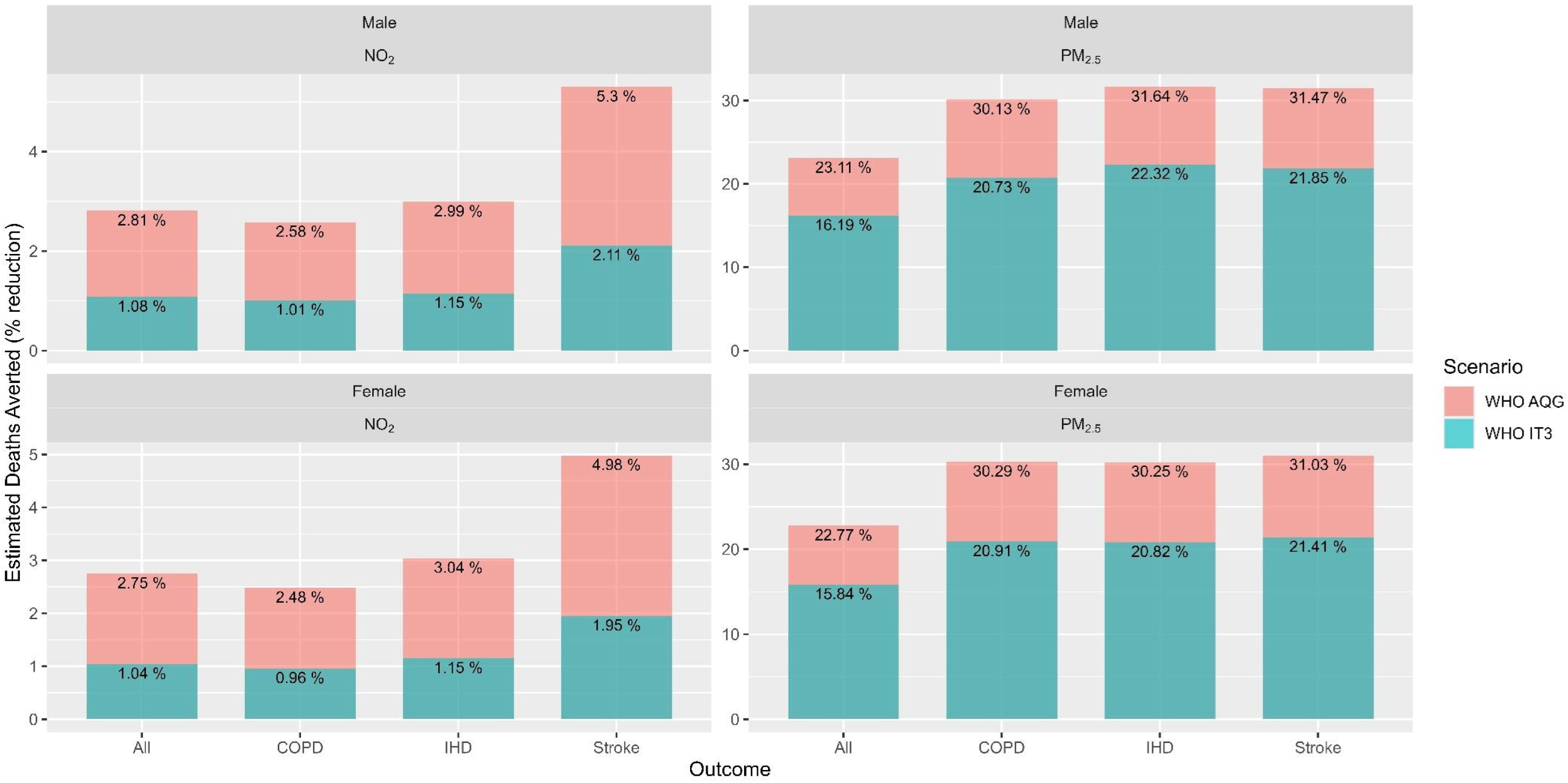
Estimated cases averted in one-year from counterfactual scenarios (WHO IT3, and WHO AQG) by continent as percentage reduction from incidence under ‘Current’ scenario (2021 IHME incidence estimates).

Absolute reductions in disease incidence increased sharply with age, particularly for IHD, stroke, and dementia, with the largest number of cases averted among individuals aged 65 years and older (Supplementary Figure 10 and 11). Across most outcomes and age groups, males experienced larger absolute reductions than females, although these differences married at older ages, reflecting higher baseline incidence among younger males.

#### Economic Costs Associated with Reduced Morbidity

COI estimates were derived for 134 countries for PM_2.5_ and 120 for NO_2_, representing market-based economic costs and do not capture intangible welfare losses. The largest estimated reductions in economic costs were associated with achieving WHO AQG levels for PM_2.5_, particularly through reductions in annual incidence of stroke (0.03% of aggregate LMIC GDP), COPD (0.026%), and IHD (0.022%) cases. Among direct medical costs, the largest estimated reductions were observed for IHD cases delayed or avoided through one year under the PM_2.5_ AQG scenario, amounting to Int$ 11.8 billion (5.3-17.2 billion). Indirect productivity-related costs were highest for COPD and stroke cases delayed or avoided under PM_2.5_ AQG scenarios, estimated at Int$ 18.1 billion (13.9-21.8 billion) and Int$ 17.9 billion (14.2-21.1 billion), respectively (Table 2). Indirect costs were not estimated for dementia, as the minimum age of incidence exceeded the assumed maximum working age, so would not capture losses associated with informal unpaid caregiving.

At the country level, China exhibited the highest combined direct and indirect economic costs associated with morbidity reductions from PM_2.5_ and NO_2_ improvements across both scenarios. For PM_2.5_, Brazil ranked number 2 for dementia incidence (direct costs only) while India ranked second for other outcomes. When expressed as a percentage of national GDP, the largest estimated economic costs were observed in Lebanon for NO_2_ reduction under the WHO AQG scenario (Supplementary Figure 12 and 13).

## Discussion

### Principal Findings

To our knowledge, few studies have jointly evaluated both mortality and morbidity outcomes, across multiple diseases, while also quantifying economic welfare and cost-of-illness impacts under WHO IT3 and AQG scenarios at this geographic scale. The findings indicate that achieving WHO air quality target levels could lead to substantial one-year reductions in mortality risk and disease incidence, particularly for cardiovascular and cerebrovascular outcomes. PM_2.5_ reductions yielded markedly larger health and economic benefits than NO_2_. Across outcomes, the largest benefits were observed for IHD and stroke, especially in countries of Africa and Asia where exposure levels and population vulnerability are greatest. Although NO_2_-related benefits were smaller in magnitude, reductions were still notable, particularly for stroke incidence and mortality. Economic valuation of these health improvements indicated large, monetised welfare benefits associated with reduced mortality risk, alongside measurable reductions in direct medical costs and productivity-related losses from reduced morbidity.

### Comparison with Existing Estimates

Global estimates of mortality attributable to ambient air pollution vary widely depending on exposure-response functions and modelling assumptions.^26^ Our estimates of PM_2.5_-related mortality reductions under WHO AQG scenarios are higher than those reported by the Health Effects Institute (4.9 million) and Global Burden of Disease studies (4.58 million), which typically rely on integrated exposure-response (IER) models.^8,27,28^ In contrast, our results align more closely with studies using Global Exposure Mortality Models (GEMM), which generally produced higher attributable mortality estimates (e.g., 7.96 million, 8.42 million, 8.8 million, 8.9 million).^6,29–31^ These differences largely reflect methodological choices. IER models incorporate evidence from lower-exposure contexts such as household air pollution and second-hand smoke, resulting in more conservative hazard ratios at higher concentrations.^23^ In contrast, this analysis relied on concentration-response functions derived from meta-analyses of outdoor air pollution studies and assumed log-linear relationships within capped exposure ranges. This approach avoids extrapolation beyond the empirical evidence base while maintaining sensitivity to marginal exposure reductions.^23^

Evidence on NO_2_-related mortality remains comparatively limited, particularly for links between long-term exposure and stroke mortality and COPD incidence, hence the use of concentration response functions that were not statistically significant (i.e., 95% confidence intervals included 1).^32,33^ Therefore findings related to these exposure-outcome pairs are indicative rather than definitive. There are few multi-country assessments on NO_2_ exposure, with available reports being focused on urban populations only.^34^ Our NO_2_-attributable mortality country-level estimates (e.g., China) are consistent with national studies and substantially higher than global urban-only assessments, underscoring the importance of rural and peri-urban exposure in LMICs.^34–36^ Differences between our results and other global assessments may also reflect variation in underlying exposure estimates. For example, national population-weighted mean NO_2_ concentrations reported in the State of Global Air and Global Burden of Disease studies are higher for several countries than those applied here, likely due to difference in modelling frameworks, including spatial resolution, satellite retrieval products, land-use regression inputs, treatment of near-road increments and population weighting approaches.^27,37^ Because NO_2_ exhibits strong intra-urban spatial heterogeneity, such methodological differences can meaningfully influence baseline exposure estimates and consequently, attributable burden calculations.^37^ For example, Song et al report zero deaths attributable to NO_2_ exposure in Ethiopia, Malaysia, Rwanda and Zambia, but this analysis estimates mortality in one year to be 2,749, 4,059, 183, and 594, respectively.^34^

Of 6.8 million deaths due to air pollution, 4.5 million are attributable to ambient PM_2.5_ and 2.1 million attributable to household air pollution (HAP).^27^ Therefore, the total burden of air pollution in LMICs is substantially underestimated when HAP is not considered. While the present analysis focuses on ambient air pollution to assess the potential benefits of achieving WHO air quality target values, effective attainment of these concentrations will likely require coordinated strategies addressing both ambient and household sources of pollution.^38,39^ As such, the benefits estimated may in fact be interpreted as potentially conservative.

### Economic Valuation of Health Impacts

Recent World Bank estimates place the global economic costs of air pollution-related mortality at approximately US$6 trillion in 2020, equivalent to 4.6% of global GDP, with a reported range of US$4.5-6.1 trillion (4.7%-6.5% of GDP).^2^ Higher welfare estimates reported here reflect key methodological differences between this analysis and World Bank’s. Here we used log-linear exposure-response functions derived from ambient air pollution studies, and income-adjusted VSLs applied to mortality risk reductions. In contrast, the World Bank reports global burden estimates using IER-based functions and valuation approaches that tend to compress VSLs in lower-income settings (i.e., valuation approaches based on uniform or low elasticities understate VSLs in lower-income contexts).^40^ Furthermore, the World Bank analysis limited mortality to only include mortality from IHD, stroke, COPD, lung cancer and lower respiratory infections, whereas here we included all-causes of mortality as defined by IHME (i.e., deaths due to all conditions).^2,41^ Whilst there may be differences in absolute values across studies, which reflect methodological choices, all studies estimate that the majority of air pollution-related economic burden arise from premature mortality rather than morbidity.^2,29^ Also consistent with these previous works, China accounted for the largest absolute share of economic welfare impacts, while other countries ranked higher when impacts were expressed relative to national GDP.^2,29^ Across studies, IHD consistently represents the largest contributor to economic welfare losses from air pollution, followed by stroke and COPD.^29^ A key contribution of this analysis is the extension of economic valuation to cause-specific morbidity and mortality for both PM_2.5_ and NO_2_, including dementia outcomes, which have rarely been monetised in prior global assessments.^27^

### Interpretation and Equity Considerations

Both VSL and COI approaches are widely used to monetise health impacts but are subject to important limitations. VSL reflects willingness to pay for marginal risk reductions and is influenced by income, risk perception, and social context, leading to systematic disparities across populations.^42–44^ Even when adjusted for GDP per capita and price levels, VSL-based estimates embed existing global income inequalities and should therefore be interpreted as descriptive valuation rather than normative measure of intrinsic human worth.^45,46^ Similarly, COI estimates capture only market-based costs and are constrained by assumptions about labour participation, wages, and productivity. As a result, they underestimate the societal burden of disease, particularly in LMICs where informal labour, unpaid caregiving, and non-market production play a substantial role.^47,48^ The absence of informal care costs, especially for conditions such as dementia, likely leads to conservative estimates of morbidity-related economic impacts.^48^ Neither VSL nor COI estimates account for the costs of implementing air quality improvements. The values reported here therefore represent economic burden and welfare implications, not net benefits or cost-effectiveness of specific interventions.

### Strengths and Implications

This study offers one of the most comprehensive assessments to date of the health and economic implications of achieving WHO air quality target values in LMICs. By jointly evaluating mortality, morbidity, welfare valuation, and cost-of-illness impacts across multiple diseases, pollutants, and scenarios, the analysis provides a nuanced picture of where air quality improvements could yield the greatest benefits, whilst disaggregating results by age, sex, and region highlights variations in vulnerability across different population groups.

Despite the alignment with global health frameworks, we acknowledge that our estimates may diverge from those reported in previous studies due to differences in methodological assumptions, data sources, and geographic focus. The findings highlight the importance of prioritising air quality interventions in highly polluted and densely populated regions, particularly in Africa and Asia, but also the need for more equitable and context-sensitive economic valuation frameworks. As countries consider air quality standards and mitigation strategies, these results reinforce the substantial societal value of cleaner air while cautioning against overinterpretation of monetised estimates as direct financial savings.

### Limitations

Firstly, reliance on openly accessible data restricted pollutant coverage to PM_2.5_ and NO_2_ and precluded assessment of pollutant mixtures or synergistic effects. The concentration-response functions applied for PM_2.5_ and NO_2_ were estimated independently and were not adjusted for co-pollutant exposure, and therefore must be interpreted independently.^49–51^ Second, while age and sex stratification was incorporated, other dimensions of vulnerability, e.g., socioeconomic status, ethnicity, and healthcare access, were not accounted for.

Furthermore, applying concentration response functions uniformly across a population assumes no differential susceptibility and that risk is the same throughout the life course, despite growing evidence contrary to this.^52^ Third, all health impacts were estimated as one-year changes and therefore reductions in mortality and morbidity represent potential delays rather than permanent prevention, and multi-morbidity interactions were not modelled.^53^ Finally, this analysis does not capture impact on quality of life, wellbeing nor impacts to wider systems, including co-benefits of emission reductions, such as climate mitigation or ecosystem impacts.

## Conclusion

Achieving WHO air quality target concentrations in LMICs could yield substantial reductions in premature mortality and disease incidence, particularly for cardiovascular, respiratory, and neurodegenerative conditions. While economic valuation highlights the large societal value of these health gains, the results should be interpreted within the limitations of customary valuation methods and the one-year, counterfactual nature of the analysis. Together, these findings support urgent and targeted air quality action while underscoring the need for equitable and comprehensive approaches to valuing human health in environmental policy.

## Data Availability

All data produced in the present study are available upon reasonable request to the authors

## Acknowledgements

The authors gratefully acknowledge Andrew Larkin, Susan Anenberg, Daniel L. Goldberg, Arash Mohegh, Michael Brauer, and Perry Hystad for providing the global nitrogen dioxide (NO_₂_) dataset used in this study. We also sincerely thank Siyuan Shen, Chi Li, Aaron van Donkelaar, Nathan Jacobs, Chenguang Wang, and Randall V. Martin for making the global PM_₂_._₅_ dataset available. Their efforts in compiling and maintaining these high-quality air pollution datasets were essential for the completion of this research.

