## Supplementary Tables and Figures for "Air Pollution, Health, and Economics: Evaluating the Impact of WHO targets and Guideline Values on Mortality and Morbidity in Low- and Middle-Income Countries"

### Methods 1. Population, pollutant and outcome selection and justification

**Population**

WorldPop’s constrained (UN adjusted) datasets presents a more accurate population distribution for urban settings than the unconstrained modelling approach.^19^ The top-down constrained estimation modelling uses satellite-derived building footprint data together with settlement maps to generate gridded population estimates at fine spatial resolution.^19^ The UN-adjusted version is scaled to align with national population totals reported by the United Nations World Population Prospects (WPP), ensuring consistency with official demographic estimates.^20^ Data is disaggregated by sex (binary only; male or female) and age categories (5 year intervals).

**Pollutants**

Global data on PM_2.5_ and NO_2_ annual average concentrations were sourced from Shen et al and Larkin et al, respectively.^22,23^ The most recent PM_2.5_ dataset available was for 2022 at a resolution of 0.01° x 0.01° (approximately 1km x 1km at the equator). The most recent NO_2_ dataset available was for 2020 at a resolution of 50m x 50m and this was aggregated to 1km x 1km. Due to limitations in computational power, disaggregating pollutant data to the population resolution was not possible and in the case of PM_2.5_ would have produced the same results. Pollutant data maps were layered over population maps, with resampling of pollutant data to the population data grid where grids did not align.

**Outcomes**

The World Health Organisation Global Air Quality Guidelines and the WHO’s Estimation of Morbidity from Air Pollution and its Economic Costs (EMAPEC) project (Table 1).^1,2^ report effects by physiological systems (e.g., cardiovascular effects), and if the disease falls within this category, we have applied the same determination of causality (e.g., IHD for cardiovascular effects). ‘Causal’ indicates the pollutant was shown to result in health effects in studies in which chance, confounding, and other biases could be ruled out with reasonable confidence. ‘Likely’ indicates the pollutant has been shown to result in adverse health effects in studies where results are not explained by chance, confounding, and other biases, but uncertainties remain in the evidence overall.^2^

Table 1: Each mortality and morbidity outcome included in the health impact assessment, along with the recommended causality determinations.^1–3^

| **Measure** | **Outcome** | **PM_2.5_** | **NO_2_** |
| --- | --- | --- | --- |
| Mortality | All-cause | Causal | Suggestive |
|  | COPD | Causal | Suggestive |
|  | IHD | Causal | Suggestive |
|  | Stroke | Causal | Suggestive |
| Incidence | COPD | Likely | Likely |
|  | Dementia | Likely | - |
|  | IHD | Causal | Suggestive |
|  | Stroke* | Causal | Suggestive |

*Grouped into the cardiovascular effects category as EMAPEC project did

### Methods 2. Economic Assessment

**Valuation of Mortality Risk**

The economic benefits of averted premature mortality were estimated using a Value of a Statistical Life (VSL) approach.^4^ A baseline VSL derived from OECD studies was first updated from 2005, estimated at Int$ 3 million (in terms of Purchasing Power Parity [PPP] USD), to 2021 value to account for changes in prices and income over time. The updated OECD VSL was then transferred to individual countries based on differences in income levels, following standard income-adjustment methods used in the literature. In this study, VSL-based estimates represent the monetised value of reductions in mortality risk, reflecting societal willingness to pay to avoid premature deaths, rather than market prices or actual expenditure.

**Time adjustment**

The OECD-based VSL estimates in 2005 was adjusted to 2021 values using consumer price inflation and real income growth:

VSL_OECD.2021_ = VSL_OECD.2005_ x ($\frac{P_{OECD.2021}}{P_{OECD.2005}}$)x ($\frac{Y_{OECD.2021}}{Y_{OECD.2005}})^{\delta}$

Where VSL_OECD.2005_ is the average VSL estimated in 2005 from a sample of OECD studies, and VSL_OECD.2021_ is the corresponding value expressed in 2021 terms. P denotes the consumer price index (CPI), and Y denotes Gross Domestic Product (GDP) per capita measured in purchasing power parity (PPP)-adjusted international dollars. The income elasticity over time ($\delta$) was assumed to be unity ($\delta$ = 1), consistent with OECD recommendations.^5^

**Spatial (Income-Transfer) adjustment**

The 2021 OECD VSL was transferred to individual countries based on relative income levels using the following expression:

VSL_k.2021_ = VSL_OECD.2021_ x ($\frac{{Y_{k2021}}}{Y_{OECD2021}}$)^ε^

Where VSL_k.2021_ is the estimated VSL for country k in 2021, Y denotes GDP per capita in PPP-adjusted international dollars, and ε is the income elasticity of VSL. An elasticity value of ε = 1.2 was applied for low- and middle-income countries, in line with existing guidance.^5^ Country-specific VSL estimates were calculated for countries where Y was available and are provided in Supplementary [Information 1](https://universityofcambridgecloud-my.sharepoint.com/:x:/r/personal/amdn2_cam_ac_uk/Documents/oda/for_submission/navaratnam_lmic_who_heia_supplementary_information_1.xlsx?d=wd2f83bbe9e554da58e8d9734e51d7ef2&csf=1&web=1&e=QF1eRg).

**Cost of Morbidity**

The economic costs associated with disease-specific morbidity were estimated using a cost-of-illness (COI) approach, including both direct medical costs and indirect productivity losses associated with each case averted through pollution reduction. Estimates were produced at the country level for each disease outcome (see Supplementary Information 1).

**Direct Medical Costs**

Direct medical costs were calculated as the project of the number of cases and the average cost of each case.

DC_k,a_ = N_k,a_ x H_k,a_

Where DC_k,a_ denotes direct medical costs for disease a in country k, N_k,a_ is the number of cases, and H_k,a_ is the average cost per case. Country-specific treatment costs for all LMICs were not consistently available from open-access databases or the academic literature. Therefore, average cost estimates were obtained from published studies and adjusted to country k using relative health expenditure levels:^6–9^

H_k,a_ = H_ref, a_ x ($\frac{T_{k}}{T_{ref}}$)

Where H_ref,a_ is the reference average costs per case for disease a, T_k_ is current health expenditure per capita (PPP-adjusted international dollars) for country k, and T_ref_ is current health expenditure per capita for the country from which the reference estimate was derived. When reference cost estimates were available from more than one country, weighted averages were calculated for reference costs and reference health expenditure using country-level incidence and population size:

| **Pooling** | **Description** |
| --- | --- |
| **Reference costs** | Reference costs were weighted by number of observations reported in each study:  H_ref, a_ = $\frac{\left( H_{A,a}\times I_{A} \right)+\left( H_{B,a}\times I_{B} \right)}{I_{A}+I_{B}}$  Where $H_{A,a}$ and $H_{B,a}$ are average cost estimates for disease a and countries A and B, respectively, and $I_{A}$ and $I_{B}$ are the corresponding numbers of observations of disease a |
| **Reference health expenditure** | Reference health expenditure per capita was calculated as a population-weighted average:  T_ref_ = $\frac{(T_{A} \times{Pop}_{A})+\left( T_{B}\times{Pop}_{B} \right)}{{Pop}_{A}+{Pop}_{B}}$  Where $T_{A}$ and $T_{B}$ denote health expenditure per capita, and ${Pop}_{A}$ and ${Pop}_{B}$ denote population sizes for the reference countries. |

Ischemic heart disease (IHD) and stroke were treated as acute conditions, with costs estimated per patient per episode. Dementia and chronic obstructive pulmonary disease (COPD) are progressive chronic conditions and therefore costs were estimated per patient per year. All cost estimates were either reported in or converted to PPP-adjusted international dollars and adjusted to the study year using the consumer price index and GDP per capita.

**Indirect Costs**

Indirect costs (i.e., productivity costs) associated with morbidity were estimated following the World Bank human capital approach, based on lost workdays due to illness:^10^

IC_k,p,a_ = D_k,p,a_ x W_k_

Where IC_k,p,a_ denotes indirect costs for disease a that is attributable to pollutant p in country k, D_k,p,a_ is the total number for disease days, and W_k_ is the average daily wage.

Total disease days were estimated as:

D_k,p,a,_ = I_k,p,a_ x d_k,a_ x t_k,a_ x 365

Where I_k,p,a_ is disease incidence, d_k,a_ is disability weight country k for disease a, and t_k,a_ is average duration of disease a in country k (in years). Disability weight, disease duration, prevalence and years lived with disability (YLD) were obtained from the Institute of Health Metrics and Evaluation (IHME).

Average daily wage was estimated as:

W_k_ = $\frac{{GDP}_{k} x S_{k}}{L_{k} x 250}$

Where GDP_k_ is gross domestic product in PPP-adjusted USD, S_k_ is the labour share of GDP, L_k_ is the total labour force, and 250 represents the average number of working days per year.

Given that indirect costs assumed labour market participation, productivity losses were capped at age 60, reflecting estimated retirement ages in several LMIC and consistent with previous analyses.^11–13^ As a result, indirect costs (i.e., productivity losses) were not estimated for dementia. These estimates of indirect costs do not account for intangible health and wellbeing losses, non-market productivity losses (e.g., unpaid household labour, informal caregiving, subsistence or informal-sector work), nor broader macroeconomic or intergenerational effects. Country-level data that was extracted or calculated can be found in Supplementary Information 1.

### Table 1. Table of OECD DAC list

| **Least**  **Developed**  **Countries** | **Low Income Countries Which Are Not LDCs (Per Capita GNI <= $1 135 In 2022)** | **Lower Middle Income Countries And Territories Which Are Not LDCs (Per Capita GNI $1 136 - $4 465 In 2022)** | **Upper Middle Income Countries And Territories Which Are Not LDCs (Per Capita GNI $4 466 - $13 845 In 2022)** |
| --- | --- | --- | --- |
| Afghanistan (L)  Angola (LM)  Bangladesh (LM)  Benin (LM)  Burkina Faso (L)  Burundi (L)  Cambodia (LM)  Central African Republic (L)  Chad (L)  Comoros (LM)  Democratic Republic of the Congo (L)  Djibouti (LM)  Eritrea (L)  Ethiopia (L)  Gambia (L)  Guinea (LM)  Guinea-Bissau (L)  Haiti (LM)  Kiribati (LM)  Lao People’s Democratic Republic (LM)  Lesotho (LM)  Liberia (L)  Madagascar (L)  Malawi (L)  Mali (L)  Mauritania (LM)  Mozambique (L)  Myanmar (LM)  Nepal (LM)  Niger (L)  Rwanda (L)  Sao Tome and Principe (LM)  Senegal (LM)  Sierra Leone (L)  Solomon Islands (LM)  Somalia (L)  South Sudan (L)  Sudan (L)  Tanzania (LM)  Timor-Leste (LM)  Togo (L)  Tuvalu (UM)  Uganda (L)  Yemen (L)  Zambia (LM | Democratic People’s Republic of Korea  Syrian Arab Republic | Algeria  Bhutan  Bolivia  Cabo Verde  Cameroon  Congo  Côte d’Ivoire  Egypt  Eswatini  Ghana  Honduras  India  Iran  Jordan  Kenya  Kyrgyzstan  Lebanon  Micronesia  Mongolia  Morocco  Nicaragua  Nigeria  Pakistan  Papua New Guinea  Philippines  Samoa  Sri Lanka  Tajikistan  Tokelau*  Tunisia  Ukraine  Uzbekistan  Vanuatu  Viet Nam  Zimbabwe | Albania  Argentina  Armenia  Azerbaijan  Belarus  Belize  Bosnia and Herzegovina  Botswana  Brazil  China (People’s Republic of)  Colombia  Costa Rica  Cuba  Dominica  Dominican Republic  Ecuador  El Salvador  Equatorial Guinea  Fiji  Gabon  Georgia  Grenada  Guatemala  Guyana2  (H)  Indonesia  Iraq  Jamaica  Kazakhstan  Kosovo  Libya  Malaysia  Maldives  Marshall Islands  Mauritius  Mexico  Moldova  Montenegro  Montserrat3  (H)  Namibia  Nauru4  (H)  Niue*  North Macedonia  Palau  Panama2  (H)  Paraguay  Peru  Saint Helena*  Saint Lucia  Saint Vincent and the Grenadines  Serbia  South Africa  Suriname  Thailand  Tonga  Türkiye  Turkmenistan  Venezuela5  Wallis and Futuna*  (the State of) Palestine |

### Table 2: Data sources

| Data type | Source | Resolution | Year |
| --- | --- | --- | --- |
| Population | WorldPop^14^ | 100m x 100m* | 2019 |
| PM_2.5_ concentration | Shen et al 2024^15^ | 1km x 1km | 2022 |
| NO_2_ concentrations | Larkin et al 2023^16^ | 50m x 50m* | 2020 |
| Health Outcomes (Incidence, Prevalence, Years of Life with Disability [YLD]) | Institute of Health Metrics^17^ | Country-level | 2021 |
| Value of Statistical Life | OECD^5^ | OECD countries | 2005 (converted to 2021) |
| Consume Price Index | World Bank Indicators^18^ | Country-level | 2005 and 2021 |
| Gross Domestic Product (GDP) per capita (PPP-adjusted Int$) | World Bank Indicators^18^ | Country-level | 2005 and 2021 |
| GDP (PPP-adjusted Int$) | World Bank Indicators^18^ | Country-level | 2021 |
| Health expenditure per capita (PPP-adjusted Int$) | World Bank Indicators^18^ | Country-level | 2021 |
| Labour Share of GDP | Our World In Data^19^ | Country-level | 2021 |
| Total Labour Force | World Bank Indicators^18^ | Country-level | 2021 |

*aggregated to 1km x 1km

### Table 3: Concentration response function (CRF) for each exposure-outcome pair used in the health impact assessment, showing risk ratios (95% Confidence Intervals) per unit increment and the upper limit of the pollutant concentration range from which the CRF was derived from (i.e., CRF derived from meta-analyses).

| **Pollutant** | **Measure** | **Outcome** | **Risk ratio (95%CI)^** | **Increment** | **Concentration upper limit** |
| --- | --- | --- | --- | --- | --- |
| **NO_2_** | Deaths | All-cause^20^ | 1.05 (1.03-1.07) | 10 **µg/m³** | 129.9 **µg/m³** |
|  |  | COPD^20^ | 1.04 (1.02-1.06) | 10 **µg/m³** | 104 **µg/m³** |
|  |  | IHD^20^ | 1.05 (1.03-1.08) | 10 **µg/m³** | 104 **µg/m³** |
|  |  | Stroke^20^ | 1.08 (0.99-1.19) | 10 **µg/m³** | 104 **µg/m³** |
|  | Incidence | COPD^21^ | 1.03 (0.91-1.16) | 10 **µg/m³** | 33.84 **µg/m³** |
|  |  | Dementia^22^ | 1.03 (1.01-1.05) | 10 **µg/m³** | 56 **µg/m³** |
|  |  | IHD^23^ | 1.01 (1.01-1.02)* | 10 µg/m³ | 94.56 **µg/m³** |
|  |  | Stroke^24^ | 1.08 (1.04-1.12) | 10 **µg/m³** | 37.78 **µg/m³** |
| **PM_2.5_** | Deaths | All-cause^25^ | 1.10 (1.06-1.13) | 10 **µg/m³** | 72.4 **µg/m³** |
|  |  | COPD^25^ | 1.14 (1.08-1.20) | 10 **µg/m³** | 43.7 **µg/m³** |
|  |  | IHD^25^ | 1.14 (1.10-1.19) | 10 **µg/m³** | 60 **µg/m³** |
|  |  | Stroke^25^ | 1.15 (1.10-1.19) | 10 **µg/m³** | 66 **µg/m³** |
|  | Incidence | COPD^26^ | 1.18 (1.13-1.23) | 10 **µg/m³** | 26 **µg/m³** |
|  |  | Dementia^22^ | 1.08 (1.02-1.14) | 5 **µg/m³** | 25 **µg/m³** |
|  |  | IHD^27^ | 1.13 (1.05-1.22) | 10 **µg/m³** | 65 **µg/m³** |
|  |  | Stroke^28^ | 1.16 (1.12-1.2) | 10 **µg/m³** | 36 **µg/m³** |

^Rounded to two decimal places; * Reported 1.022 (1.016-1.029) 10ppb increment, converted to 10µg/m³

### Table 4: One-year change in all-cause and cause-specific mortality under counterfactual air pollution reduction scenarios (WHO IT3 and WHO AQG), disaggregated by sex (male and female), and associated VSL-based economic welfare estimates of deaths averted, expressed relative to aggregate LMIC GDP. Total GDP for NO_2_ is Int$ 78,100.84 and PM_2.5_ Int$78,142.4 bill.

| Sex | Pollutant | Outcome | Scenario | Total population (billions) | Total Deaths in 'Current' Scenario 2021 (thousands) | Estimated Deaths Averted (thousands) | Rate of Estimated Deaths Averted (per 100,000) | Estimated Deaths Averted (% reduction) | VSL-based Estimated costs of Deaths Averted (Int$ billions) | VSL-based Estimated costs of Deaths Averted (% of GDP) |
| --- | --- | --- | --- | --- | --- | --- | --- | --- | --- | --- |
| Male | NO_2_ | All | WHO IT3 | 1.48 | 23498604 | 253 (156-345) | 17.11 | 1.08 | 340 (209-463) | 0.44 |
|  |  |  | WHO AQG | 1.48 | 23498604 | 661 (408-901) | 44.76 | 2.81 | 859 (530-1170) | 1.1 |
|  |  | COPD | WHO IT3 | 1.48 | 1594741 | 16 (8-23) | 1.09 | 1.01 | 23 (12-34) | 0.03 |
|  |  |  | WHO AQG | 1.48 | 1594741 | 41 (21-60) | 2.78 | 2.58 | 57 (29-83) | 0.07 |
|  |  | IHD | WHO IT3 | 1.48 | 3473536 | 40 (25-61) | 2.69 | 1.15 | 54 (33-83) | 0.07 |
|  |  |  | WHO AQG | 1.48 | 3473536 | 104 (64-160) | 7.03 | 2.99 | 137 (84-210) | 0.17 |
|  |  | Stroke | WHO IT3 | 1.48 | 2809111 | 59 (-8-123) | 4.02 | 2.11 | 88 (-12-183) | 0.11 |
|  |  |  | WHO AQG | 1.48 | 2809111 | 149 (-21-308) | 10.08 | 5.3 | 216 (-31-446) | 0.28 |
|  | PM_2.5_ | All | WHO IT3 | 1.48 | 23507421 | 3806 (2726-4790) | 257.56 | 16.19 | 3197 (2278-4046) | 4.09 |
|  |  |  | WHO AQG | 1.48 | 23507421 | 5434 (3919-6795) | 367.74 | 23.11 | 4846 (3476-6092) | 6.2 |
|  |  | COPD | WHO IT3 | 1.48 | 1595049 | 331 (209-437) | 22.38 | 20.73 | 327 (206-433) | 0.42 |
|  |  |  | WHO AQG | 1.48 | 1595049 | 481 (309-624) | 32.52 | 30.13 | 493 (316-642) | 0.63 |
|  |  | IHD | WHO IT3 | 1.48 | 3474894 | 776 (594-940) | 52.5 | 22.32 | 695 (530-847) | 0.89 |
|  |  |  | WHO AQG | 1.48 | 3474894 | 1100 (850-1321) | 74.42 | 31.64 | 1046 (804-1262) | 1.34 |
|  |  | Stroke | WHO IT3 | 1.48 | 2810020 | 614 (458-751) | 41.55 | 21.85 | 670 (497-823) | 0.86 |
|  |  |  | WHO AQG | 1.48 | 2810020 | 884 (667-1071) | 59.85 | 31.47 | 998 (749-1214) | 1.28 |
| Female | NO_2_ | All | WHO IT3 | 1.49 | 17534411 | 183 (113-249) | 12.28 | 1.04 | 246 (152-336) | 0.32 |
|  |  |  | WHO AQG | 1.49 | 17534411 | 483 (298-658) | 32.46 | 2.75 | 627 (387-854) | 0.8 |
|  |  | COPD | WHO IT3 | 1.49 | 1163924 | 11 (6-16) | 0.75 | 0.96 | 16 (8-23) | 0.02 |
|  |  |  | WHO AQG | 1.49 | 1163924 | 29 (15-42) | 1.94 | 2.48 | 39 (20-57) | 0.05 |
|  |  | IHD | WHO IT3 | 1.49 | 2625874 | 30 (19-46) | 2.02 | 1.15 | 42 (26-64) | 0.05 |
|  |  |  | WHO AQG | 1.49 | 2625874 | 80 (49-123) | 5.37 | 3.04 | 108 (67-166) | 0.14 |
|  |  | Stroke | WHO IT3 | 1.49 | 2327814 | 45 (-6-94) | 3.05 | 1.95 | 66 (-9-137) | 0.08 |
|  |  |  | WHO AQG | 1.49 | 2327814 | 116 (-16-240) | 7.79 | 4.98 | 164 (-23-339) | 0.21 |
|  | PM_2.5_ | All | WHO IT3 | 1.49 | 17542029 | 2778 (1990-3498) | 186.67 | 15.84 | 2296 (1636-2905) | 2.94 |
|  |  |  | WHO AQG | 1.49 | 17542029 | 3995 (2880-4998) | 268.41 | 22.77 | 3519 (2523-4425) | 4.5 |
|  |  | COPD | WHO IT3 | 1.49 | 1164134 | 243 (154-322) | 16.35 | 20.91 | 230 (145-304) | 0.29 |
|  |  |  | WHO AQG | 1.49 | 1164134 | 353 (227-458) | 23.69 | 30.29 | 345 (221-449) | 0.44 |
|  |  | IHD | WHO IT3 | 1.49 | 2626910 | 547 (419-664) | 36.75 | 20.82 | 511 (389-622) | 0.65 |
|  |  |  | WHO AQG | 1.49 | 2626910 | 795 (614-956) | 53.39 | 30.25 | 790 (607-956) | 1.01 |
|  |  | Stroke | WHO IT3 | 1.49 | 2328625 | 498 (372-609) | 33.49 | 21.41 | 512 (381-629) | 0.66 |
|  |  |  | WHO AQG | 1.49 | 2328625 | 722 (545-875) | 48.54 | 31.03 | 773 (580-940) | 0.99 |

### Figure 1: Flow chart of data exclusion of each stage of the analysis, with the number in brackets representing the number of LMICs.

There were no world bank data for Niue and Tokelau, and no GDP data for Democratic Republic of Korea. Additionally, there was no GDP per capita data for these countries and Venezuela, so VSL could not be calculated for these four countries, and no PPP data (reported or conversion factor) was available for Cuba, Eritrea, South Sudan or Yemen. All other countries’ GDP and GDP per capita data was from 2021. Direct costs for COPD data (China, Kyrgyz Republic, Mexico and Thailand), were reported in US market value, so had to be converted using Price Level Ratio. Direct cost calculations for three countries were not possible as no Health Expenditure data was available for Democratic Republic of Korea, Niue, Tokelau. It was not possible to calculate indirect costs for 12 countries because of missing labour share data (Albania), both missing labour share and force data (Dominica, Grenada, Kiribati, Marshall Islands, Micronesia (Federated States of), Nauru, Niue, Palau, Tokelau, Tuvalu), or missing GDP data (Democratic Republic of Korea).

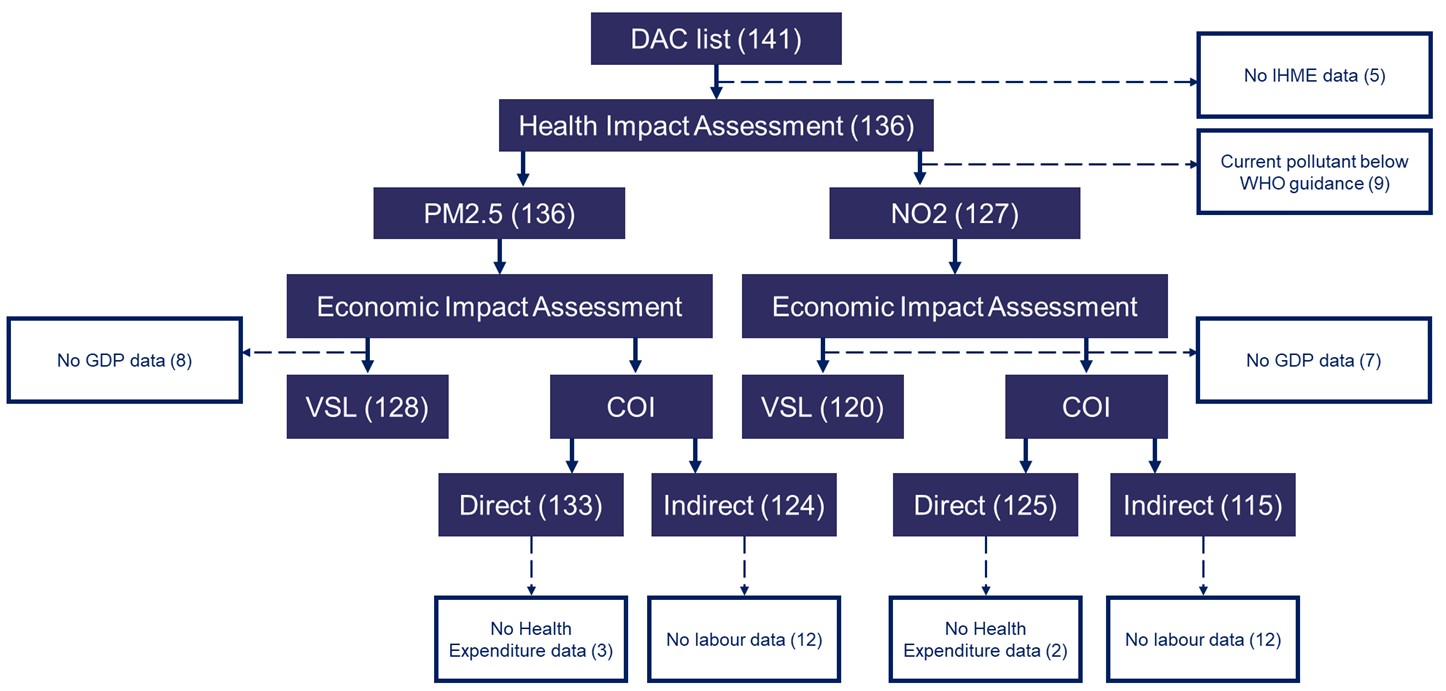

### Figure 2: Twenty countries ranked by relative (%) reductions in estimated deaths averted by PM_2.5_ concentration reduction under WHO IT3 and AQG scenarios, by outcome.

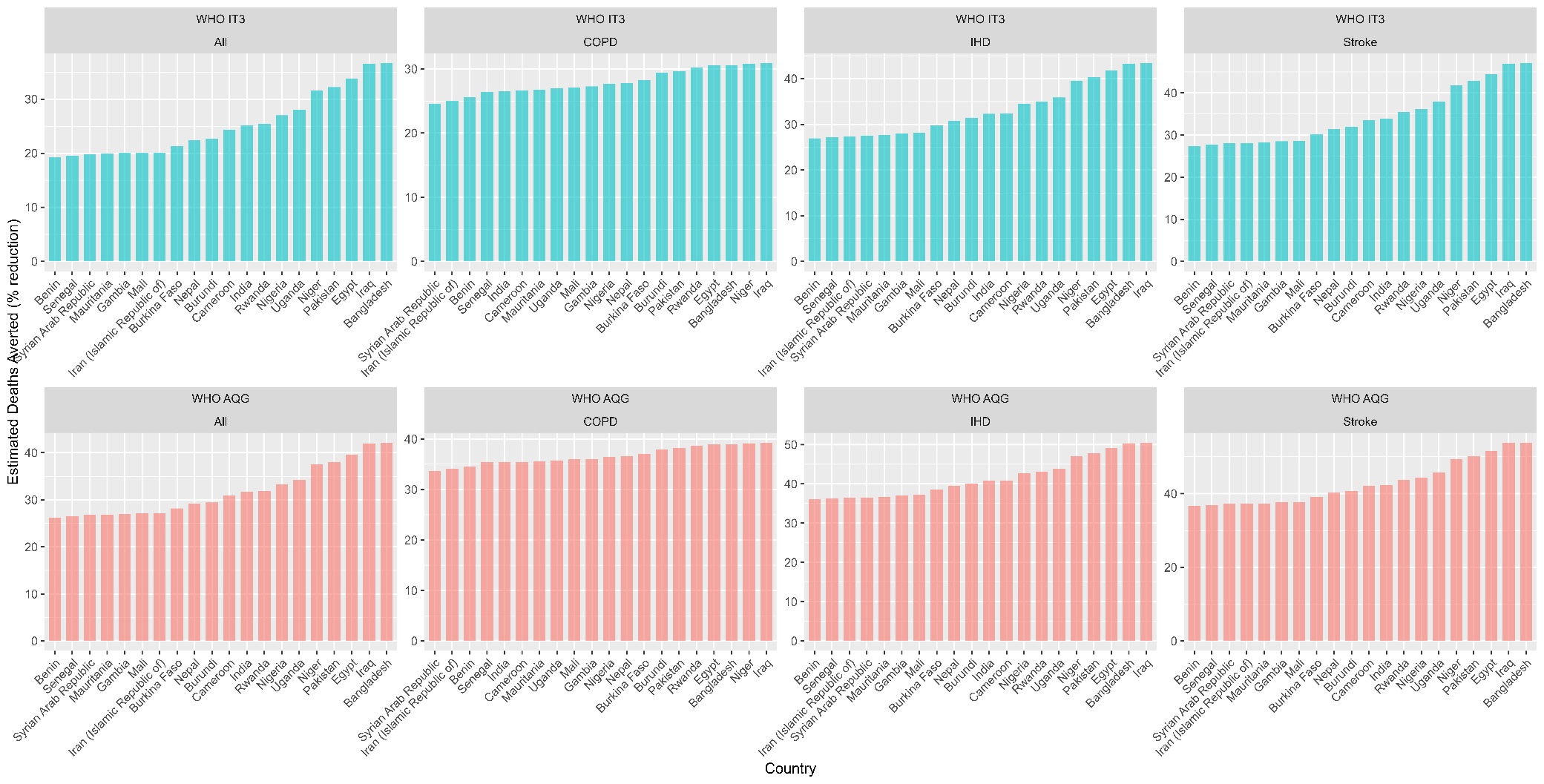

### Figure 3: Twenty countries ranked by relative (%) reductions in estimated deaths averted by NO_2_ concentration reduction under WHO IT3 and AQG scenarios, by outcome.

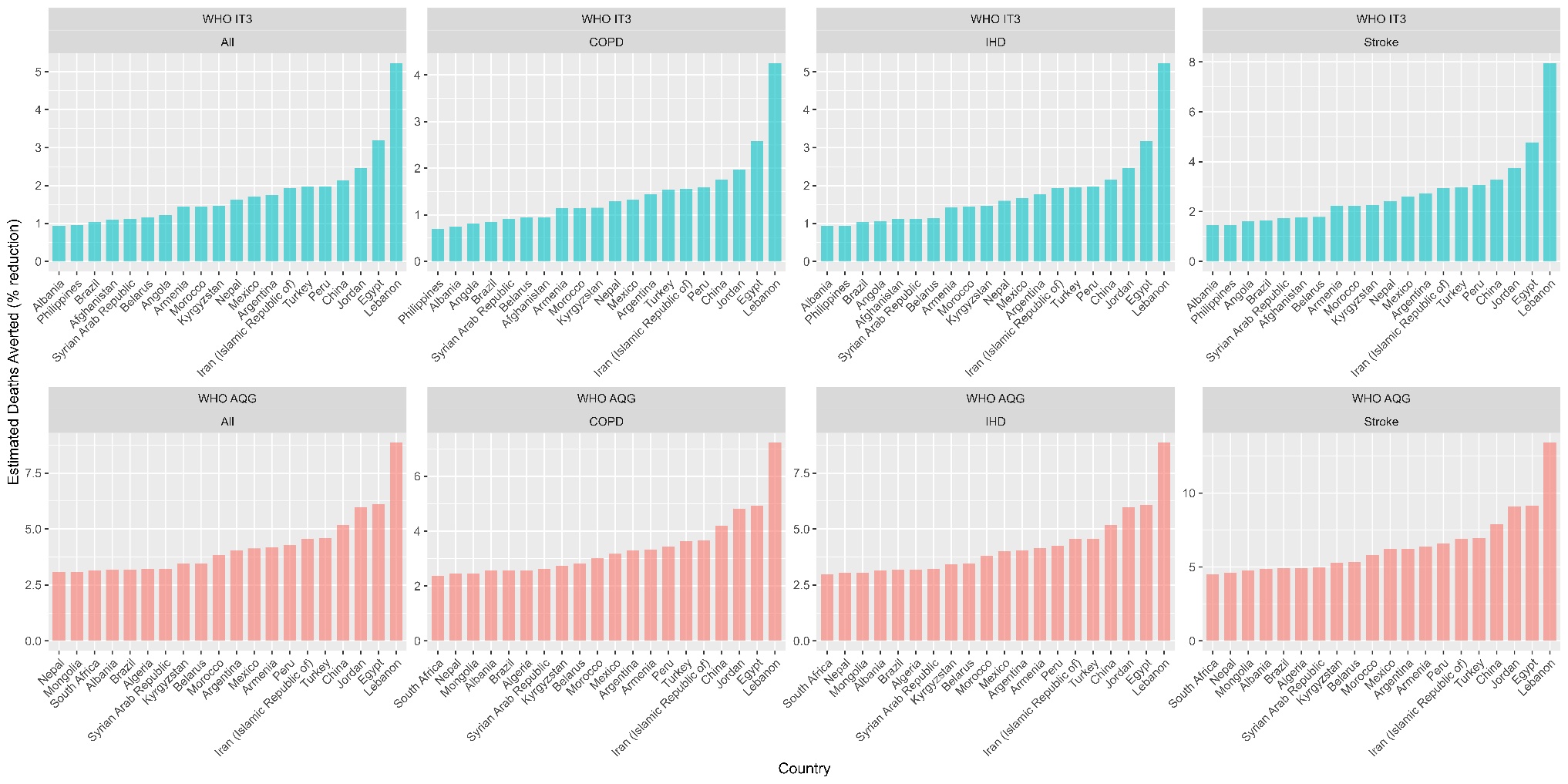

### Figure 4: One-year change in all-cause and cause-specific mortality under counterfactual air pollution reduction scenarios (WHO IT3), disaggregated by sex (male and female) and age categories (5-year increments, apart from 80, which includes all ages from 80 years onwards).

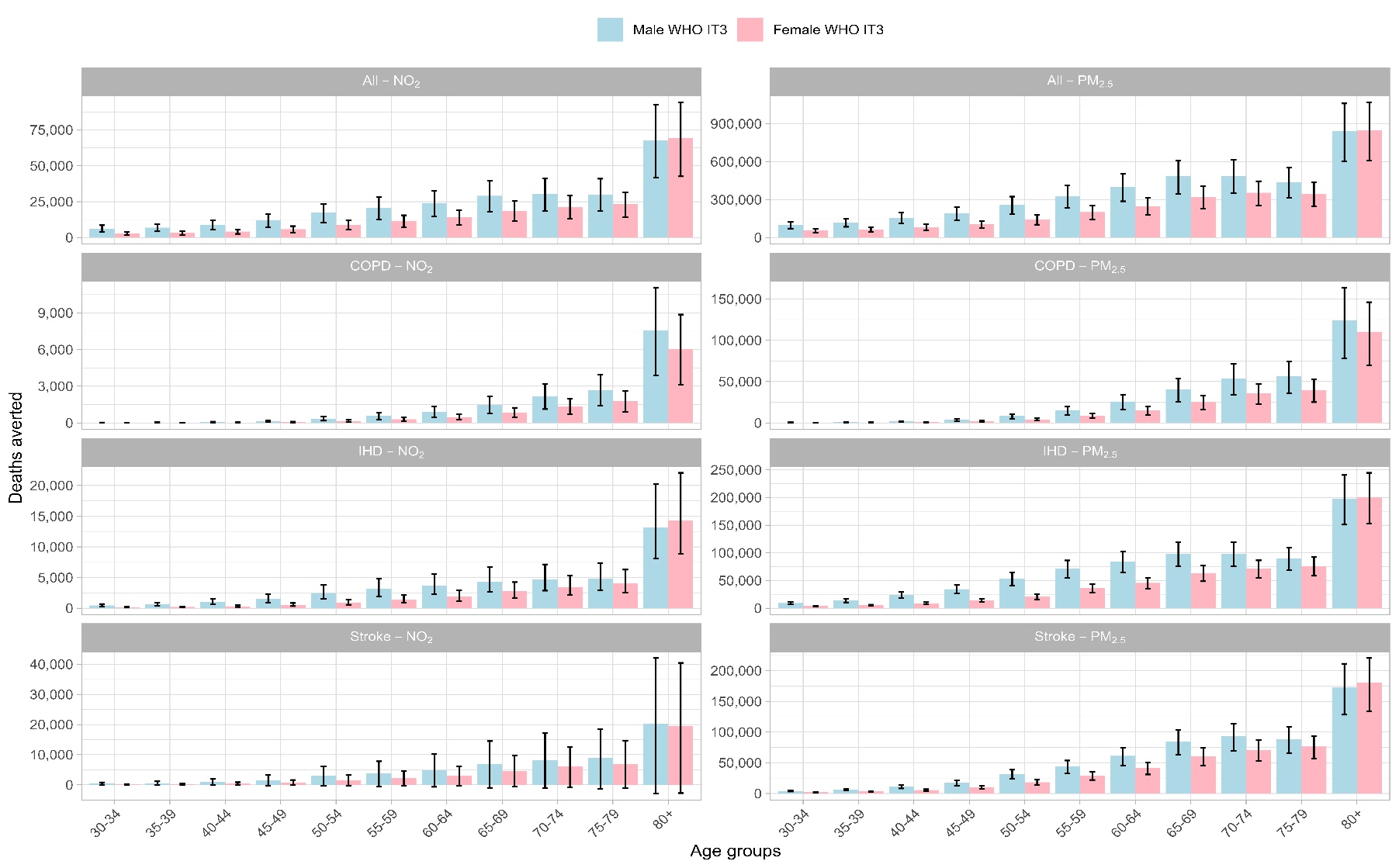

### Figure 5: One-year change in all-cause and cause-specific mortality under counterfactual air pollution reduction scenarios (WHO AQG), disaggregated by sex (male and female) and age categories (5-year increments, apart from 80, which includes all ages from 80 years onwards).

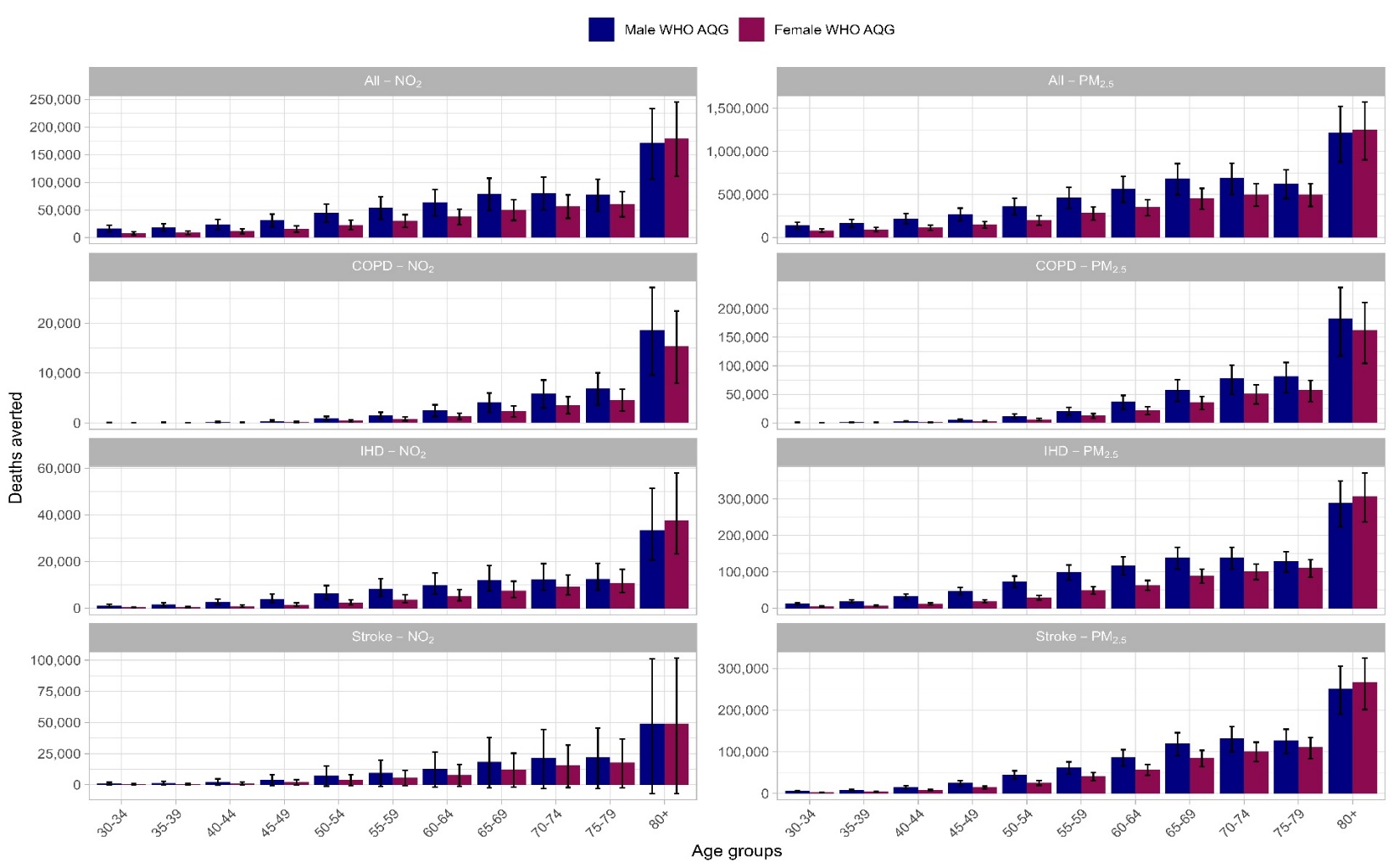

### Figure 6: Twenty countries ranked by relative (%) reductions in VSL-based monetary value of values deaths averted by PM_2.5_ concentration reduction under WHO IT3 and AQG scenarios, by outcome.

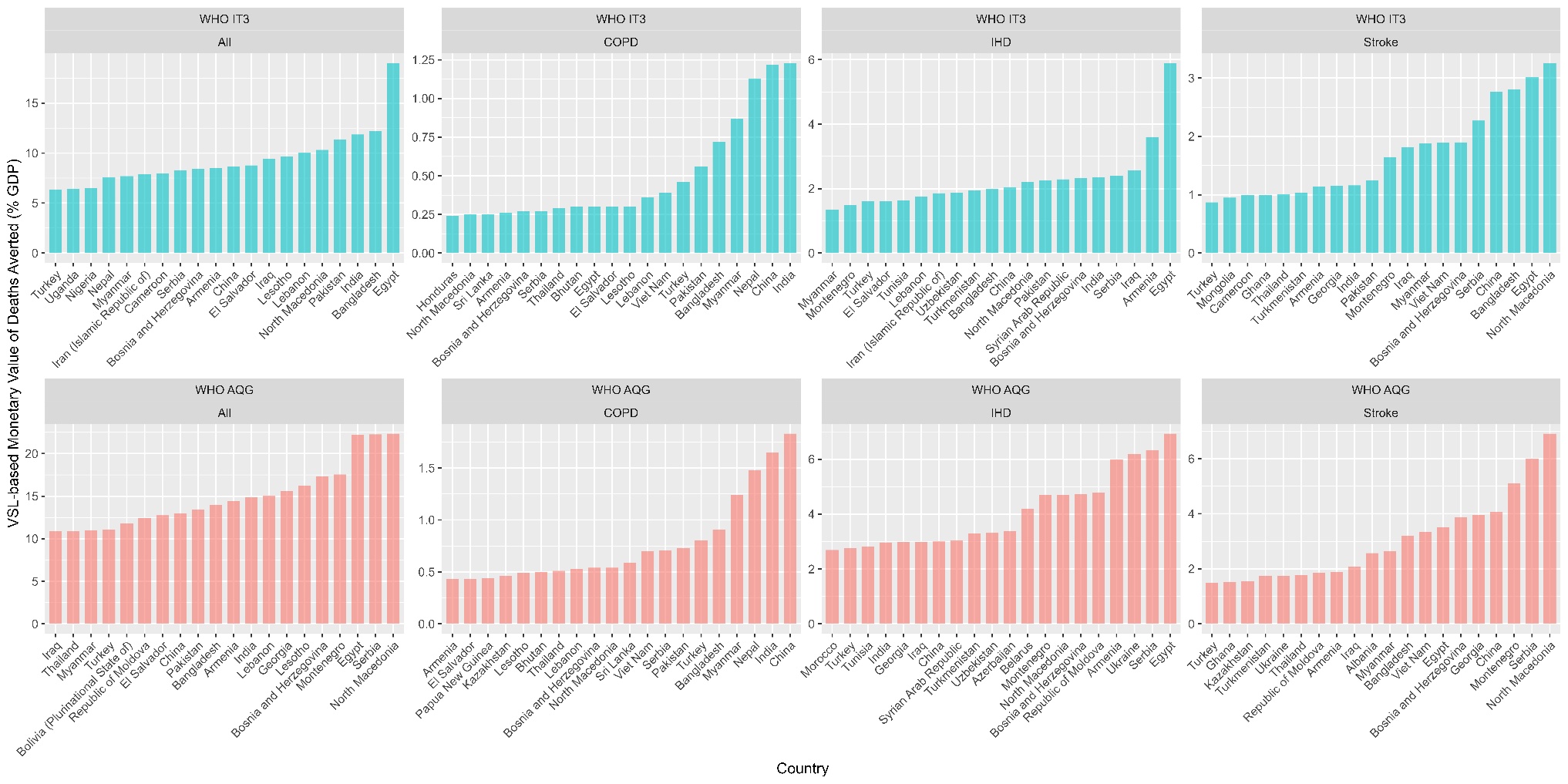

### Figure 7: Twenty countries ranked by relative (%) reductions in VSL-based monetary value of values deaths averted by NO_2_ concentration reduction under WHO IT3 and AQG scenarios, by outcome.

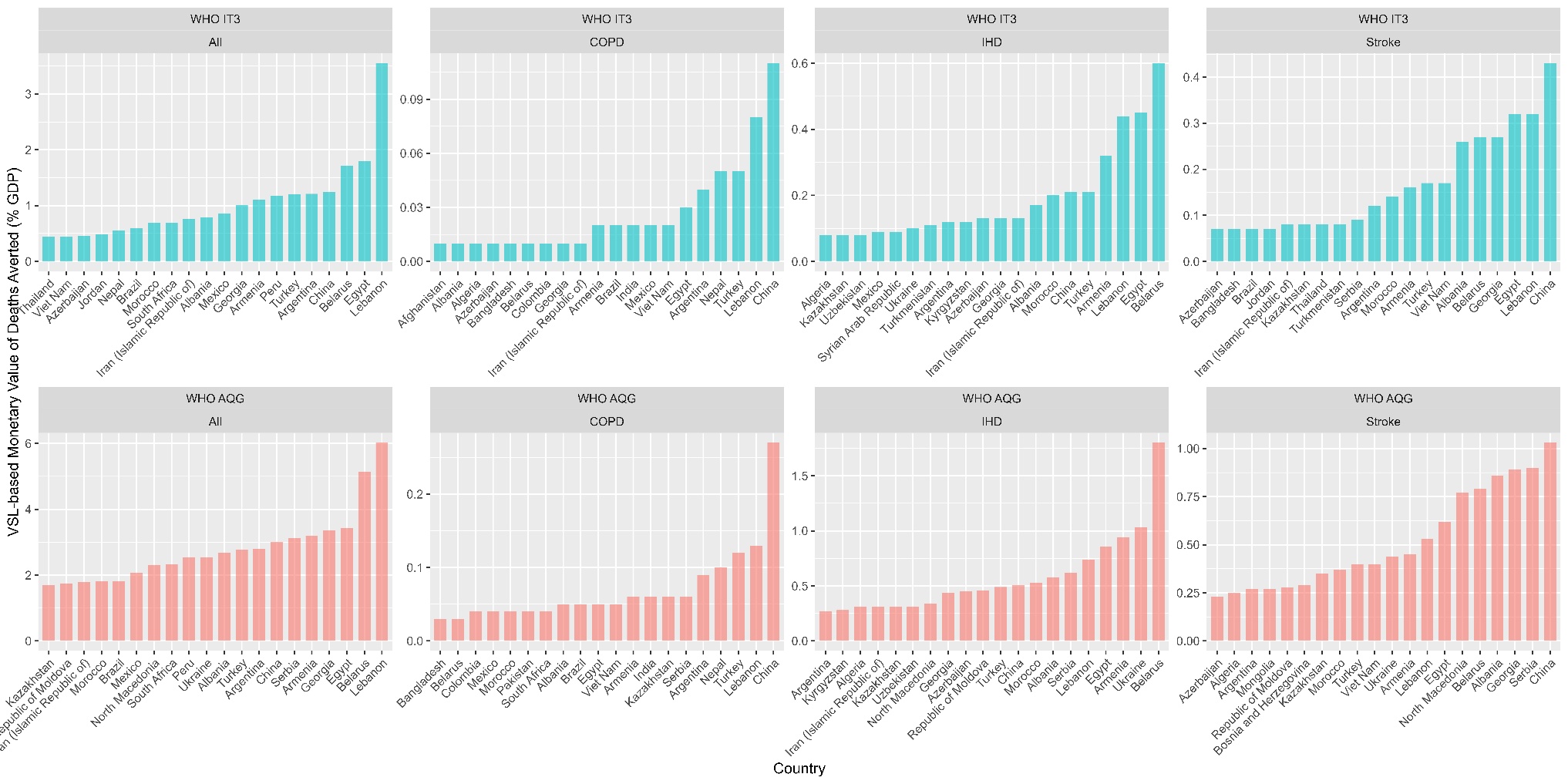

### Figure 8: Twenty countries ranked by relative (%) reductions in cases by PM_2.5_ concentration reduction under WHO IT3 and AQG scenarios, by outcome.

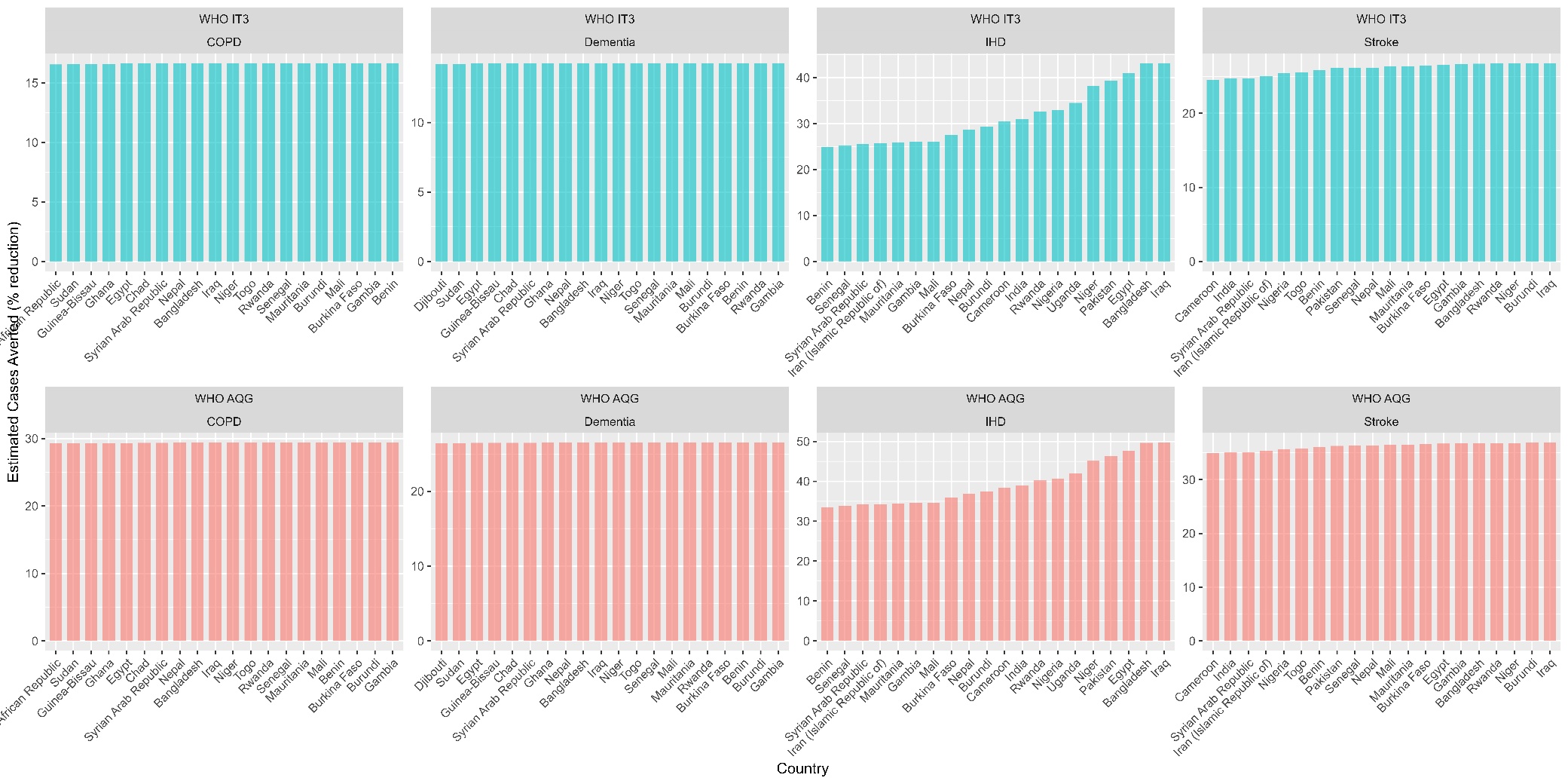

### Figure 9: Twenty countries ranked by relative (%) reductions in cases by NO_2_ concentration reduction under WHO IT3 and AQG scenarios, by outcome.

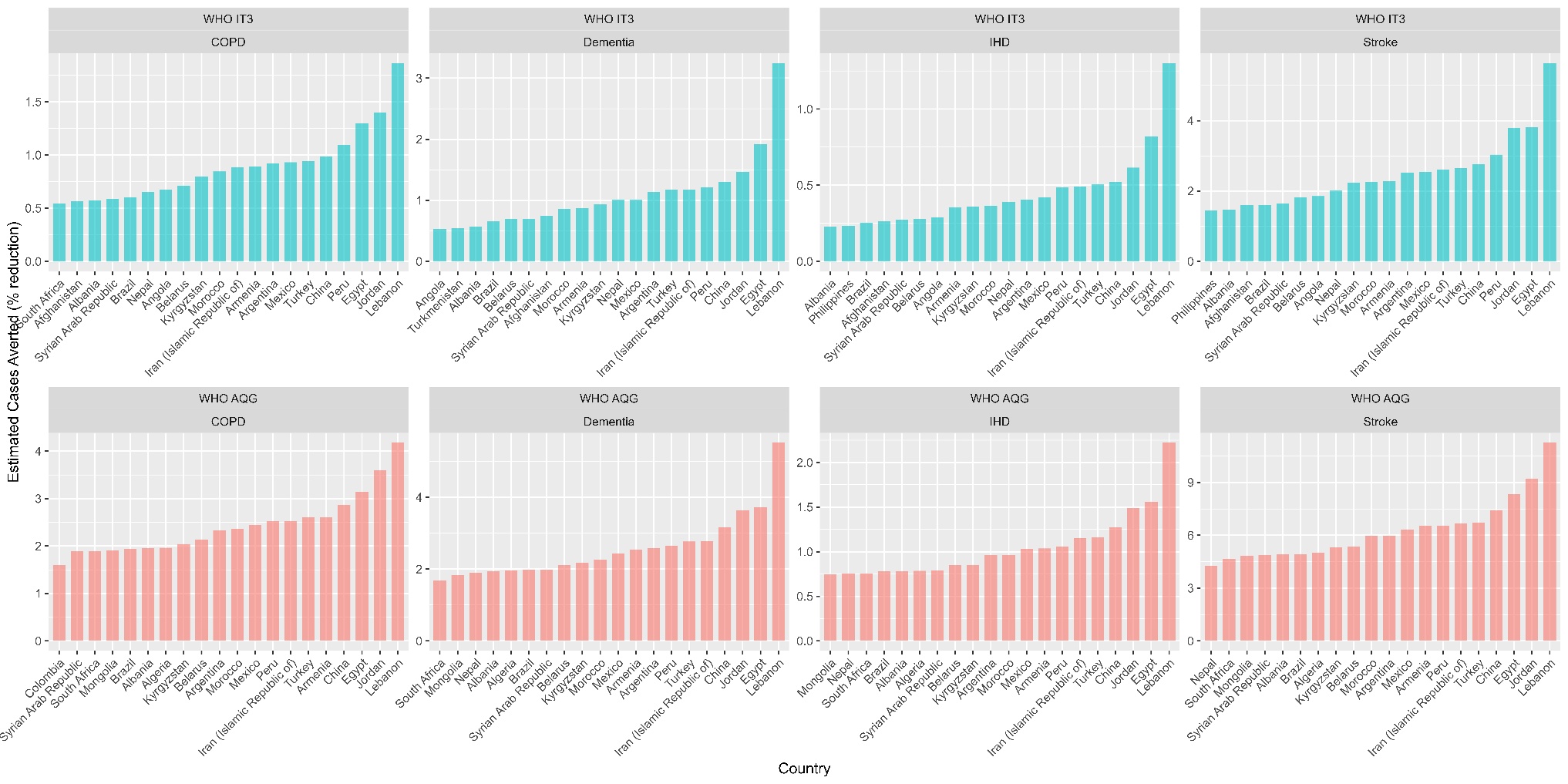

### Figure 10: One-year change in disease incidence under counterfactual air pollution reduction scenarios (WHO IT3), disaggregated by sex (male and female) and age categories (5-year increments, apart from 80, which includes all ages from 80 years onwards).

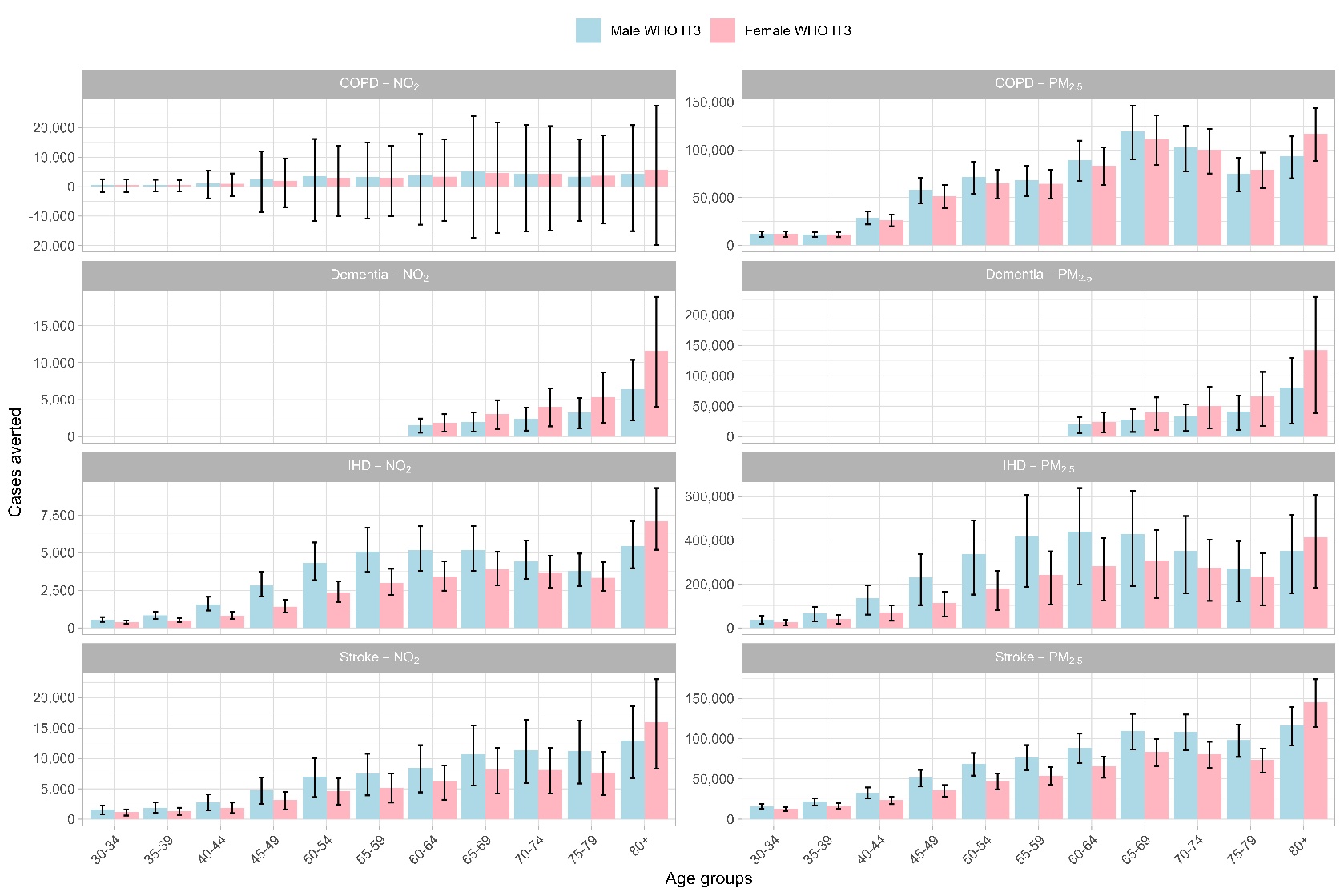

### Figure 11: One-year change in disease incidence under counterfactual air pollution reduction scenarios (WHO AQG), disaggregated by sex (male and female) and age categories (5-year increments, apart from 80, which includes all ages from 80 years onwards).

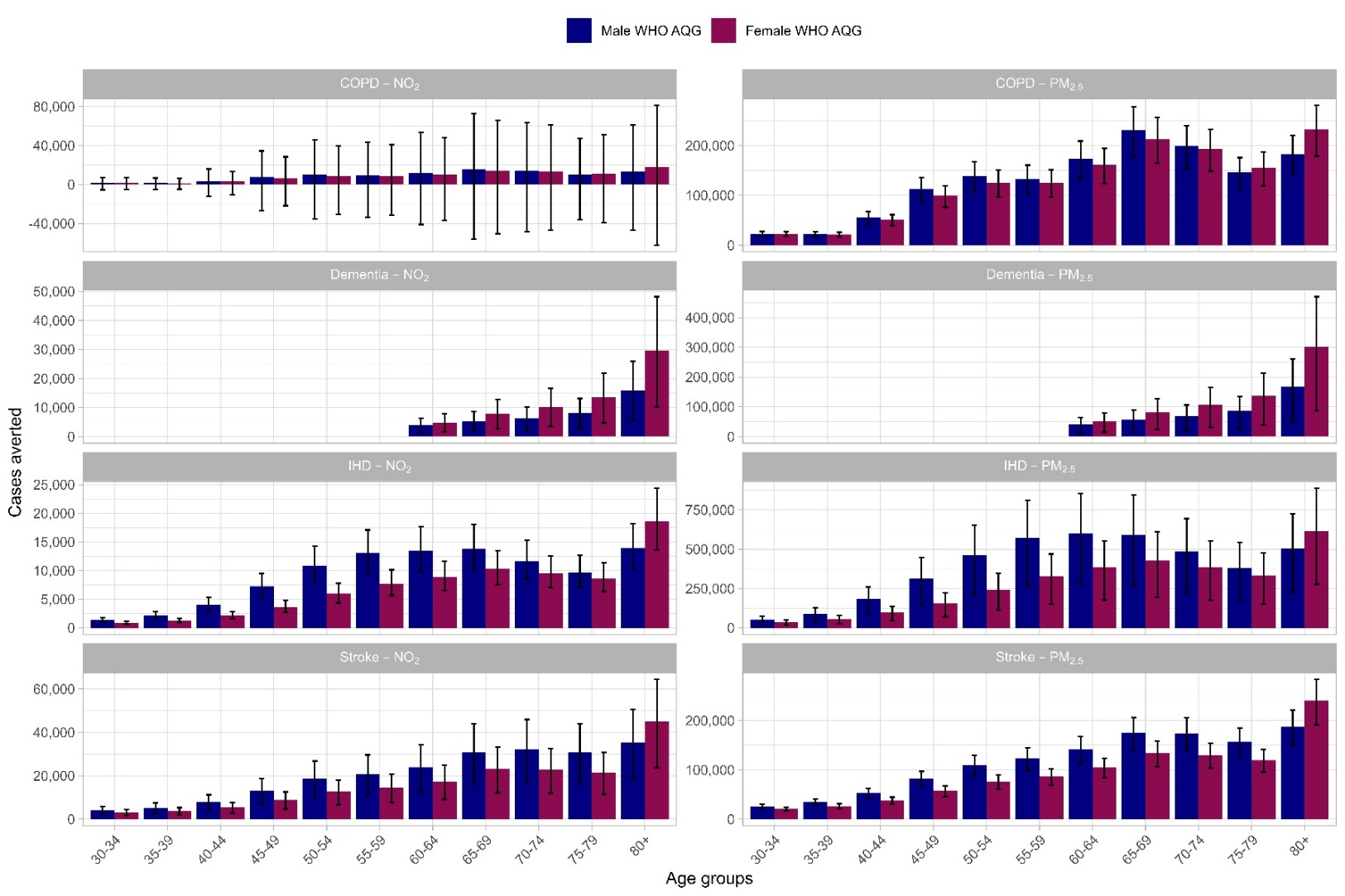

### Figure 12: Twenty countries ranked by relative (%) reductions in Cost-of Illness (direct medical and indirect productivity costs) averted by PM_2.5_ concentration reduction under scenarios WHO IT3 and AQG scenarios by outcome.

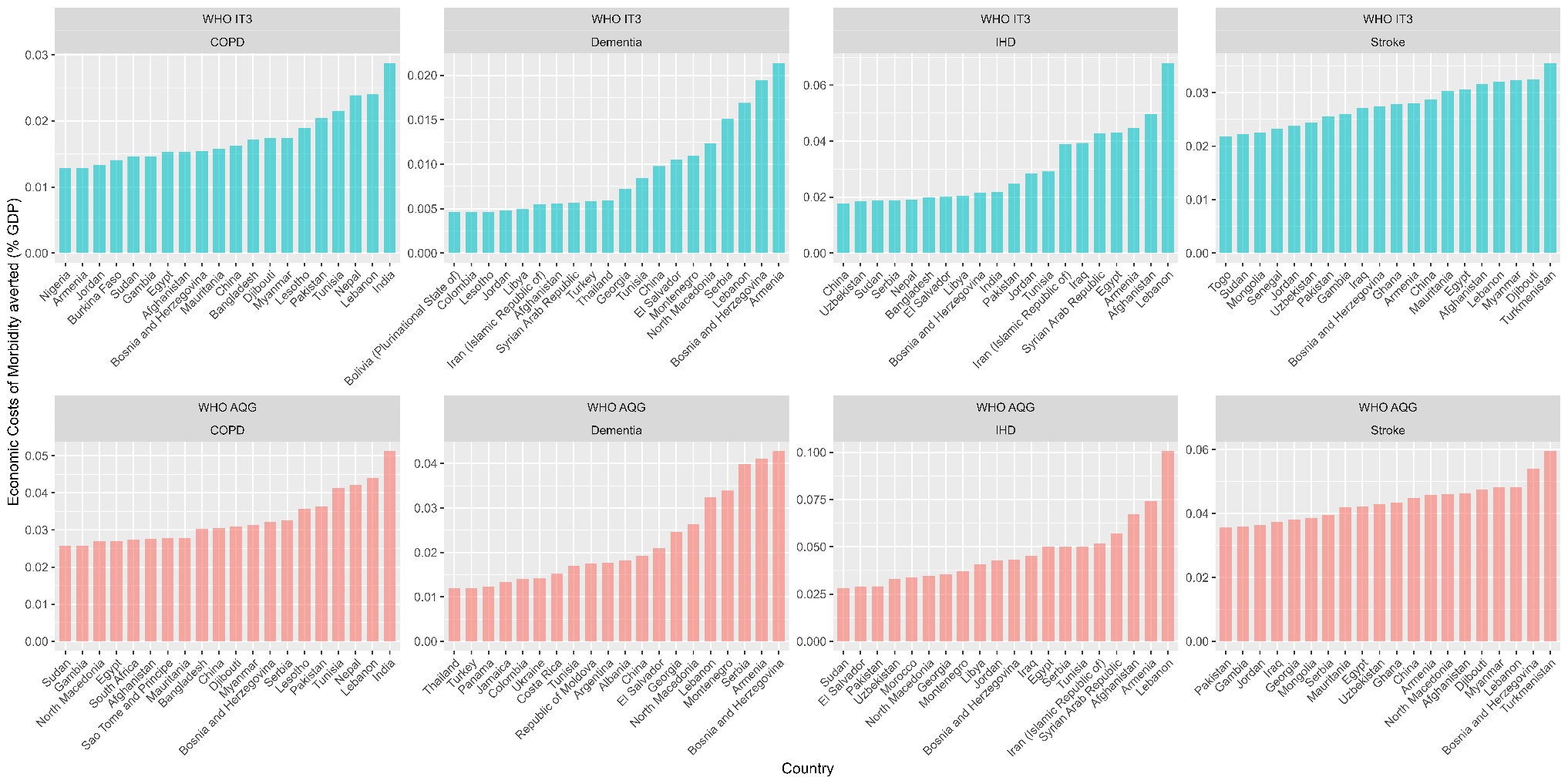

### Figure 13: Twenty countries ranked by relative (%) reductions in Cost-of Illness (direct medical and indirect productivity costs) averted by NO_2_ concentration reduction under scenarios WHO IT3 and AQG scenarios by outcome.

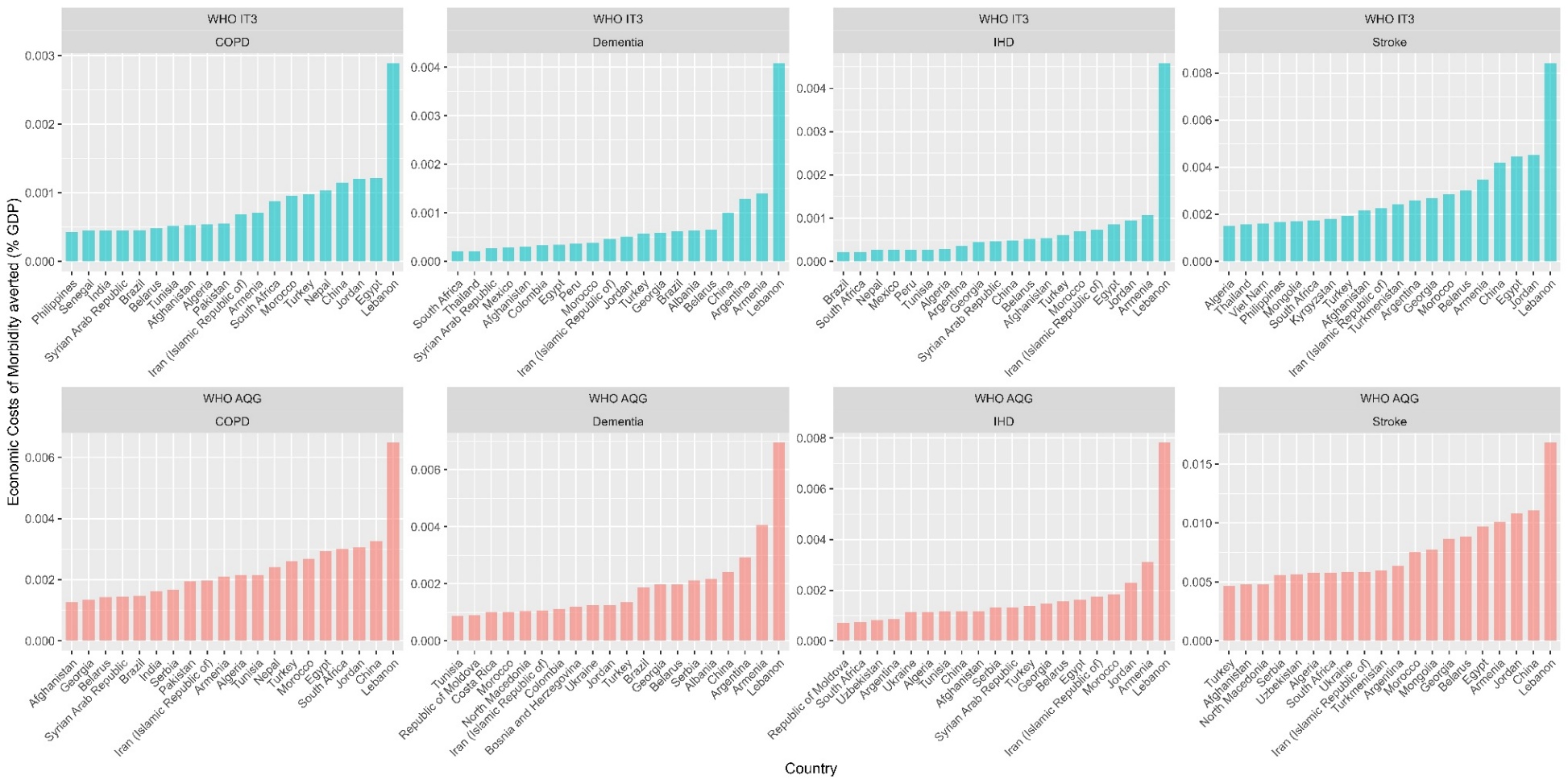

21	Haverkate TMI, Charalampous P, Gonzato E, Pereda A, Garcia V, Brauer M, Devleesschauwer B, Breitner-Busch S, Haagsma JA. Long-term ambient nitrogen dioxide exposure and COPD: a systematic review and quantitative synthesis. *Submitted*.

22	Best Rogowski CB, Bredell C, Shi Y, *et al.* Long-term air pollution exposure and incident dementia: a systematic review and meta-analysis. *Lancet Planet Health* 2025; **9**: 101266.

23	Stieb DM, Zheng C, Salama D, *et al.* Systematic review and meta-analysis of case-crossover and time-series studies of short term outdoor nitrogen dioxide exposure and ischemic heart disease morbidity. *Environ Health* 2020; **19**: 47.

24	Wolf K, Hoffmann B, Andersen ZJ, *et al.* Long-term exposure to low-level ambient air pollution and incidence of stroke and coronary heart disease: a pooled analysis of six European cohorts within the ELAPSE project. *Lancet Planet Health* 2021; **5**: e620–32.

25	Orellano P, Kasdagli M-I, Pérez Velasco R, Samoli E. Long-Term Exposure to Particulate Matter and Mortality: An Update of the WHO Global Air Quality Guidelines Systematic Review and Meta-Analysis. *Int J Public Health* 2024; **69**: 1607683.

26	Park J, Kim H-J, Lee C-H, Lee CH, Lee HW. Impact of long-term exposure to ambient air pollution on the incidence of chronic obstructive pulmonary disease: A systematic review and meta-analysis. *Environmental Research* 2021; **194**: 110703.

27	Zhu W, Cai J, Hu Y, *et al.* Long-term exposure to fine particulate matter relates with incident myocardial infarction (MI) risks and post-MI mortality: A meta-analysis. *Chemosphere* 2021; **267**: 128903.

28	Yuan S, Wang J, Jiang Q, *et al.* Long-term exposure to PM2.5 and stroke: A systematic review and meta-analysis of cohort studies. *Environ Res* 2019; **177**: 108587.
